# D4Z4End2End: complete genetic and epigenetic architecture of D4Z4 macrosatellites in FSHD, BAMS and reference cohorts

**DOI:** 10.1101/2025.04.24.25326320

**Authors:** Lucinda C. Xiao, Ayush Semwal, Brianna St John, Kathleen Zeglinski, Shian Su, James Lancaster, Shifeng Xue, Bruno Reversade, Matthew E. Ritchie, Frédérique Magdinier, Marnie E. Blewitt, Quentin Gouil

## Abstract

The D4Z4 locus is a macrosatellite array on chromosome 4q that normally comprises 8 to >100 3.3-kb repeat units. Its size and repetitiveness render it refractory to most sequencing technologies; consequently its genetic and epigenetic architectures remain incompletely understood despite their relevance to human health, in particular facioscapulohumeral muscular dystrophy (FSHD). Molecular diagnosis for FSHD following clinical description currently involves complex, multi-step and low resolution assays, aiming at identifying contractions on permissive haplotypes (FSHD type 1) or epigenetic reactivation (FSHD type 2) due to pathogenic variants in the epigenetic machinery (most often in *SMCHD1*). Here we leverage ultra-long whole-genome and Cas9-targeted sequencing to develop a fast and accurate workflow, D4Z4End2End, to comprehensively charactere the genetics and methylation of D4Z4 alleles. We apply it to samples from patients affected by FSHD1, FSHD2, and another disease caused by *SMCHD1* variants, Bosma arhinia microphthalmia syndrome (BAMS), as well as publicly-available data from the 1000 Genomes Project and Human Pangenome Reference Consortium. We attain high read depth sequencing of full-length D4Z4 arrays of up to 40 repeat units (~132 kb), accurately capture contracted arrays, genetic mosaicism, and pathogenic *SMCHD1* variants, and generate accurate consensus sequences of the full set of D4Z4 alleles for variant analysis. Moreover, we identify new allelic variants, analyse complex D4Z4 rearrangements including in-*cis* duplications, and reveal striking length- and *SMCHD1* status-dependent methylation patterns across the D4Z4 array. Our findings provide new insights into human macrosatellite genetics and epigenetics, and demonstrate the potential of long-read nanopore sequencing to accelerate FSHD research and diagnostics.

## Introduction

Facioscapulohumeral muscular dystrophy (FSHD) is a genetic disorder that causes progressive weakness of the muscles of the face, scapula and upper arms, as well as the lower legs, hip girdle and abdomen (1, 2). It is the third most common muscular dystrophy worldwide, with an estimated prevalence of between 1/8000-20000 (3–5).

FSHD has a complex aetiology involving genetic and epigenetic dysregulation of the D4Z4 macrosatellite array in the subtelomere of chromosome 4 (4q35). The D4Z4 array comprises 1 to >100 3.3-kb tandem repeats, each of which contains a partial copy of the *DUX4* gene, with a full-length *DUX4* gene being present at the distal end of the array (Figure 1A). *DUX4* encodes a transcription factor that is normally expressed in a small window of embryonic development, where it plays a key role in zygotic genome activation (6), but is thereafter silenced in most somatic tissues (7). FSHD arises when loss of silencing at D4Z4 leads to aberrant expression of *DUX4* in skeletal muscle, and subsequent activation of myotoxic signalling pathways (8).

**Fig. 1.**
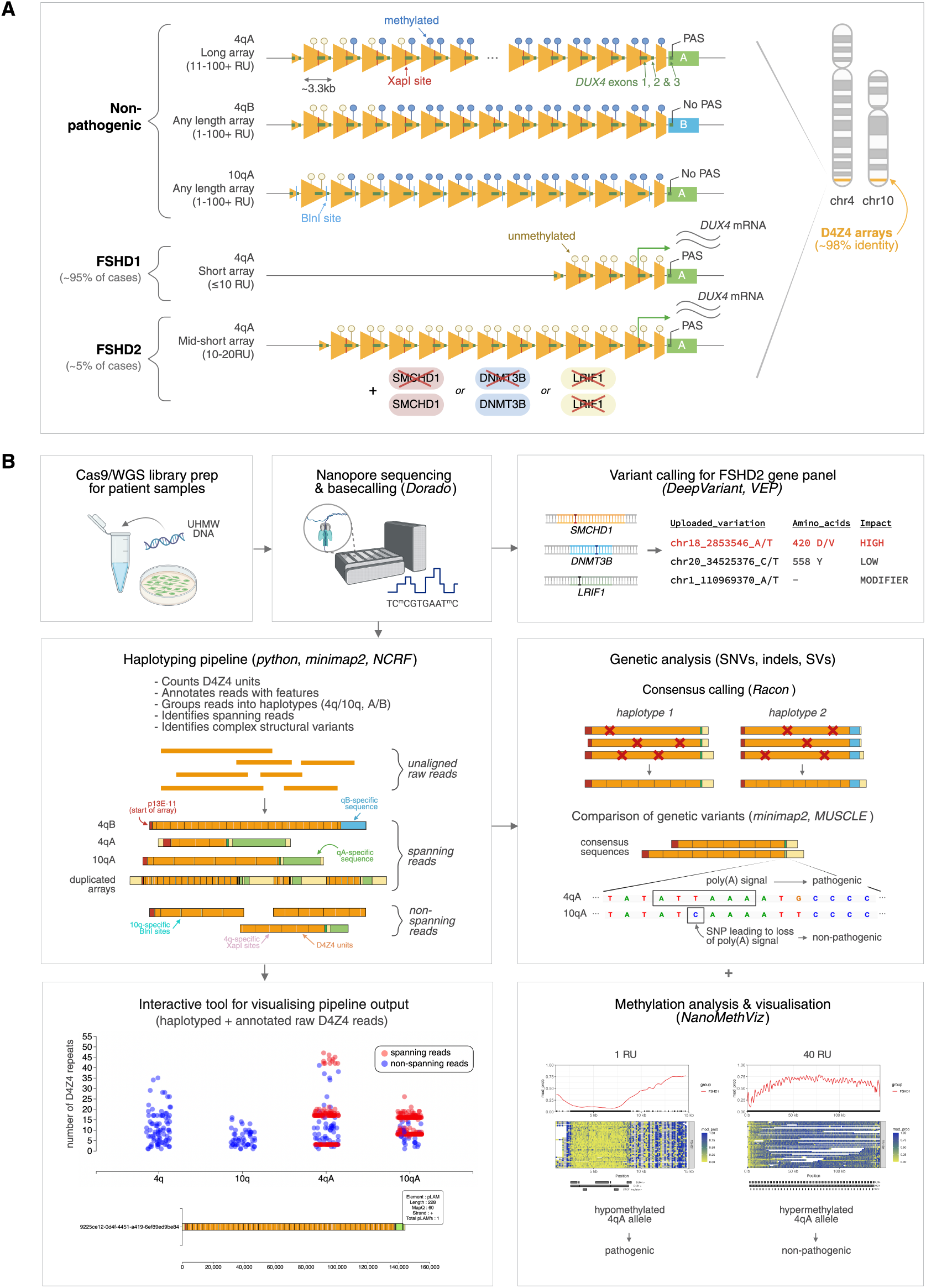
D4Z4End2End: an all-in-one, long-read-based workflow for the genetic and epigenetic analysis of FSHD. (A) Schematic of the pathogenesis of FSHD. The causative locus of FSHD is the D4Z4 macrosatellite array at the chromosome 4q subtelomere, which comprises 1 to >100 *~*3.3 kb tandem repeat units. FSHD occurs in the presence of i) a polyadenylation signal (PAS)-containing D4Z4 haplotype (4qA), which enables stable *DUX4* expression and ii) epigenetic dysregulation of the array, which can occur via array contraction (FSHD1) or pathogenic variants in chromatin modifiers (FSHD2). Both FSHD1 and FSHD2 are associated with D4Z4 hypomethylation and ectopic *DUX4* expression in skeletal muscle. A non-pathogenic, homologous D4Z4 array is located at the chromosome 10q subtelomere. A 4q-specific XapI site and 10q-specific BlnI site is used to distinguish 4q and 10q alleles in Southern blot assays. (B) Overview of the all-in-one, long-read-based workflow for FSHD. Ultra-high-molecular-weight (UHMW) DNA is used to perform Nanopore Cas9-targeted or whole-genome sequencing (WGS). Canonical and 5mCG basecalling is performed using Dorado. Variants in reads overlapping FSHD2-associated genes are called using DeepVariant (23), and the potential pathogenicity of these variants is assessed using Variant Effect Predictor (VEP) (24). For assessment of the D4Z4 array, raw reads are input into a custom script which annotates and haplotypes D4Z4 reads. The output of the haplotyping pipeline can be explored using an interactive visualisation tool, which displays raw reads as individual dots grouped into their haplotypes and plotted based on their number of D4Z4 repeat units (y-axis), and highlights 4q and 10q spanning reads. Hovering over individual dots displays the annotated raw read. Accurate consensus sequences for 4q and 10q alleles can be generated from spanning reads using Racon (25), enabling analysis of allele-specific D4Z4 genetic variants. Allele-specific, array-wide methylation analysis is performed using NanoMethViz (26, 27), allowing detection and visualisation of FSHD1- and FSHD2-specific methylation patterns.

Epigenetic derepression of D4Z4 can occur through two different mechanisms, which define the two main subtypes of FSHD (FSHD1 and FSHD2) (Figure 1A) (9). FSHD1 accounts for the majority of cases (~95%), and is caused by contraction of the D4Z4 array to 1-10 repeat units, which can either be inherited in an autosomal dominant manner or arise *de novo* (10–30% of cases) (10, 11). FSHD2 (~5% of cases) is caused by pathogenic variants in chromatin modifiers including *SMCHD1* (12), *DNMT3B* (13) and *LRIF1* (14) in the presence of an intermediate-length array of commonly 8–20 repeat units (15) or *cis*-duplicated arrays with short distal arrays (16). Non-contracted D4Z4 arrays generally have high levels of DNA methylation, whereas both FSHD1 and FSHD2 are associated with hypomethylation of either the contracted array (FSHD1) or all D4Z4 alleles (FSHD2) (2, 17).

Further factors complicate the genetics of FSHD (Figure 1A). Two main haplotypes of the 4q D4Z4 allele have been identified, designated 4qA and 4qB, based on differences in the sequence immediately distal to the D4Z4 array (18). Only the 4qA haplotype contains a polyadenylation signal (PAS) required for stabilisation of the *DUX4* mRNA, and thus only 4qA alleles are pathogenic (9). Moreover, there is a homologous qA-type D4Z4 array on chromosome 10q that shares 98% identity with the 4q D4Z4 array (19, 20), yet 10qA alleles contain a single nucleotide variant (SNV) within the PAS that renders them non-pathogenic (9). Further D4Z4 subhaplotypes have been identified based on other genetic variants within and adjacent to the D4Z4 array (21), including two variants of the 4qA allele (4qAS and 4qAL) that differ in the lengths of their distal D4Z4 units, but have been found to be equally pathogenic (22). These factors should be accounted for to provide accurate diagnoses for FSHD.

While previous studies have done much to advance our understanding of FSHD, many aspects of D4Z4 genetics and epigenetics remain poorly understood. It is still unclear the exact mechanisms by which epigenetic factors normally interact with D4Z4 repeats to mediate silencing of the array. Moreover, even amongst patients with the same number of D4Z4 repeats, there is great variability in onset, severity and penetrance of FSHD (28–30), suggesting the existence of as-yet unresolved modifiers of the disease. A prominent example of this is the ‘gray zone’ in the repeat number threshold for FSHD1, where 4qA alleles with 8-10 repeat units can give rise to FSHD, but are also found in unaffected control individuals (15). Additionally, Bosma arhinia microphthalmia syndrome (BAMS), another phenotypically-distinct human disease caused by pathogenic *SMCHD1* variants, has also been associated with D4Z4 hypomethylation (31), yet most BAMS patients do not have the skeletal muscle manifestations of FSHD (32, 33). These outstanding questions all highlight the need for methods that can more comprehensively characterise the genetics and epigenetics of D4Z4 regulation.

Until recently, study of D4Z4 has been limited by the low resolution and/or low read length of prior techniques. This is reflected in the current ‘gold standard’ for FSHD diagnostics, which after clinical assessment minimally includes counting of D4Z4 repeat number via Southern blotting to detect pathogenic contractions that are consistent with FSHD1 (15). The Southern blotting assay is able to distinguish 4q/10q alleles based on haplotype-specific SNVs that create a 4q-specific XapI restriction enzyme site, and a 10q-specific BlnI restriction enzyme site, however provides no further information about the 4q haplotype or epigenetic status. Therefore, if this first test shows that the patient does not have a 4q allele of 1-7 repeat units (strongly suggestive of FSHD), additional assays may be required to determine A/B haplotype (additional Southern blotting assay), evaluate FSHD2-associated genes (exome sequencing), or assess for D4Z4 hypomethylation (bisulfite sequencing) (15). Using this traditional diagnostic approach, it can take months to years for a patient to receive a definitive FSHD diagnosis. Moreover, more complex cases involving mosaicism and complex D4Z4 alleles, including rare 4qA haplotypes, D4Z4 duplications, hybrid arrays, D4Z4 proximal extended deletion (DPED) alleles, and 4q/10q translocations, can give rise to false positives or false negatives, or are often unable to be resolved (15).

Newer technologies such as molecular combing and optical genome mapping have enabled more in-depth characterisation of complex D4Z4 alleles that are missed by traditional technologies (34–36). Nevertheless, these assays are still limited by their inability to resolve the base-level D4Z4 sequence, and do not provide methylation information. Several groups have also developed bisulfite-sequencing-based methylation assays that have been shown to accurately diagnose FSHD1 and FSHD2 (37–39), but can still only provide the average methylation across all D4Z4 units and/or target the final D4Z4 repeat, limiting the study of array-wide D4Z4 methylation patterns.

More recently, several studies have shown the potential of long-read Nanopore sequencing for the study of the D4Z4 macrosatellite array (40–46). In this study, we extend upon these results (Supplementary Table S1) by developing an all-in-one, long-read-based workflow for the genetic and epigenetic analysis of FSHD (D4Z4End2End). D4Z4End2End combines high-read-depth Cas9-targeted sequencing of 4q and 10q D4Z4 arrays with a custom script for annotation and haplotyping of raw D4Z4 reads, alongside targeted sequencing and variant calling for a panel of FSHD2-associated genes (Figure 1B). Protocol optimisations allow for the capture of genetic mosaicism and full-length arrays of up to 40 repeat units (~142 kb including flanking regions). We combine this targeted assay with whole-genome ultra-long sequencing to reveal potential biases and complex genetic rearrangements that may be missed with a targeted approach.

We apply D4Z4End2End to the study of several FSHD1, FSHD2, BAMS and control samples, and demonstrate its ability to resolve the genetics and methylation of complete sets of patient 4q and 10q alleles. Accurate haplotyping enables us to perform in-depth genetic analysis of D4Z4 alleles, identify several previously-unreported D4Z4 variants, and document a variety of unique DNA methylation patterns across full-length D4Z4 arrays in both health and disease. Moreover, for the first time we characterise the base-level genetic structure and methylation of full-length D4Z4 duplication and triplication alleles, providing further clues towards the (epi)genetic machinery involved in the genesis and regulation of these complex D4Z4 rearrangements. These novel findings show that targeted long-read nanopore sequencing can serve as a powerful tool for the study of macrosatellite repeats, and for more informative and streamlined FSHD diagnostics.

## Results

### Haplotyping and annotation of 4q/10q long-read sequencing data to characterise D4Z4 alleles

We developed a pipeline to process and visualise long-read sequencing data covering the 4q35 and 10q26 regions, including assignment to chromosome 4q/10q, determination of A/B haplotype, counting of D4Z4 repeat units, and highlighting of reads that span the entire D4Z4 array (Figure 1B). Reads that overlap the D4Z4 array were identified by aligning the raw reads against the D4Z4 sequence and the 4q and 10q reference sequences from CHM13v2.0 (47). XapI and BlnI restriction sites, used to identify each allele in current diagnostic approaches, were also assessed within the raw reads, to confirm correct assignment to 4q or 10q. We further assigned the reads to A and B haplotypes based on the presence of pLAM (qA-specific sequence) and qB-specific sequences, respectively. Reads that contained both p13E-11 (a region flanking the proximal end of the D4Z4 array) and either pLAM or qB-specific sequence were classified as spanning reads, and these spanning reads were used to determine the haplotypes and number of D4Z4 units for each allele. To aid in visualisation of the results, we created an interactive tool that can be used to explore individual raw 4q and 10q reads that have been processed by the pipeline and annotated with their features (p13E-11, D4Z4 units, pLAM, qB-specific sequence, and XapI and BlnI restriction sites) and haplotypes (4q/10q and A/B) (Figure 1B, Figure 2).

**Fig. 2.**
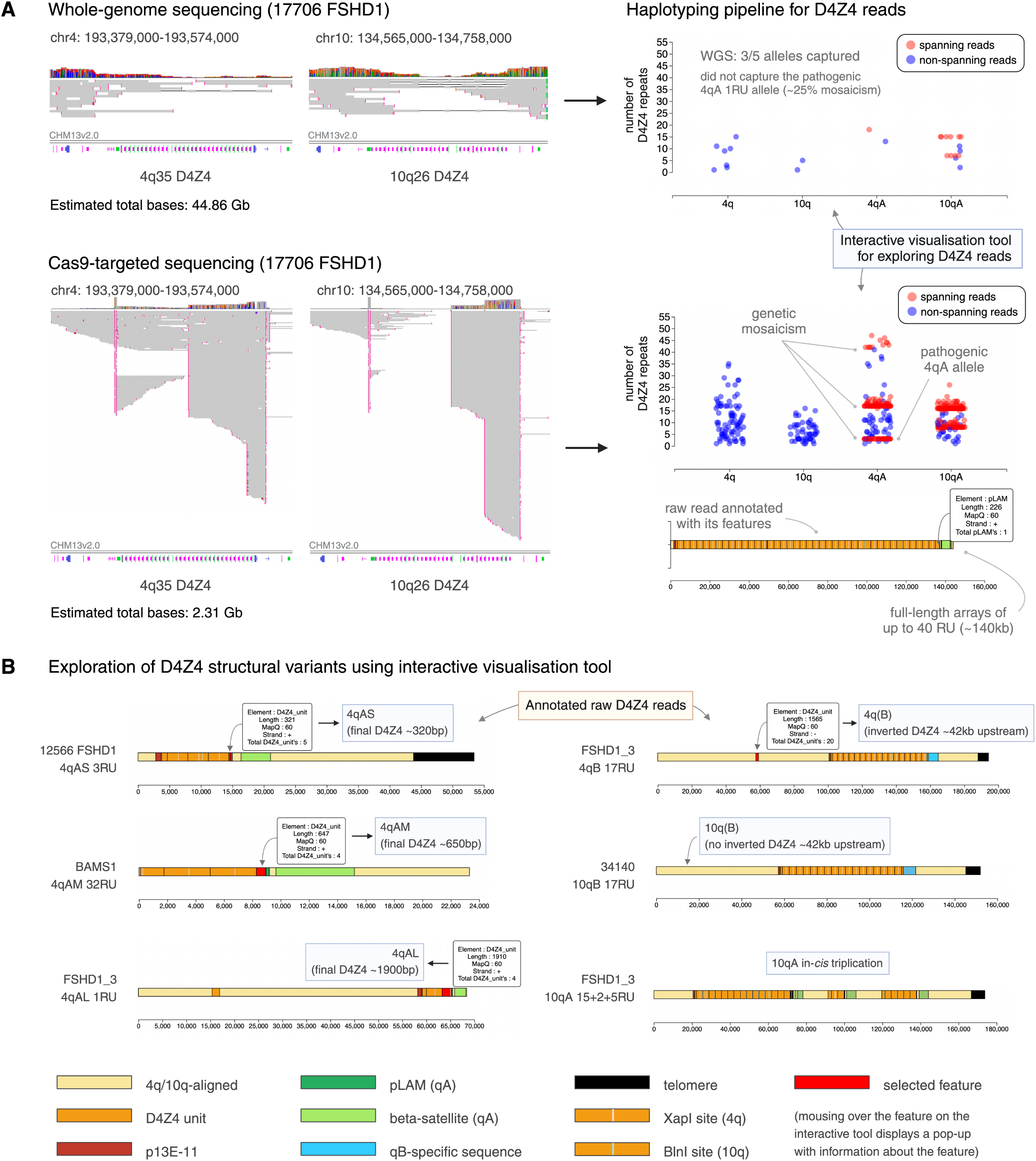
An interactive tool for exploring annotated and haplotyped D4Z4 raw reads from long-read sequencing data. (A) Sequencing data obtained for the 4q35 and 10q26 D4Z4 regions using ultra-long whole-genome sequencing (WGS) and Cas9-targeted sequencing, for sample 17706 (FSHD1). (Left) Alignment of sequencing data for the D4Z4 region against a single reference genome, such as CHM13v2.0 (32 repeat units for both the 4q35 and 10q26 D4Z4 arrays), leads to large gaps in alignment and potential mis-mapping of reads, making interpretation difficult. (Right) Our pipeline processes the sequencing data to annotate raw reads with their D4Z4 features, enabling accurate repeat unit counting and assignment to 4q/10q and A/B haplotypes. (Top) WGS generates a high total sequencing output (44.86 Gb for sample 17706) but yields low coverage of the 4q35 and 10q26 regions, which can lead to pathogenic alleles being missed. (Bottom) Cas9-targeted sequencing leads to much higher coverage of the 4q35 and 10q26 regions with a much lower total sequencing output (2.31 Gb for sample 17706), which enables the capture of low-frequency pathogenic alleles, genetic mosaicism, long full-length arrays, and high numbers of spanning reads. (B) The pipeline is accompanied by an interactive visualisation tool that enables exploration of the structure of raw D4Z4 reads, revealing D4Z4 variants such as (Left) 4qAS, 4qAL and a newly-identified 4qAM allele, as well as (Right) 10qB alleles from 4q/10q translocations, and in-cis triplication alleles.

We first used the pipeline to analyse publicly-available raw Nanopore data and assess assemblies published by the Human Pangenome Reference Consortium (HPRC) (48), which have previously been studied in the context of FSHD (49). The pipeline was able to successfully identify and annotate raw reads that spanned complete D4Z4 arrays, enabling accurate D4Z4 counting and resolution of 4q and 10q alleles (Supplementary Table S2). Despite the presence of raw spanning reads in the Nanopore data, assessment of the associated HPRC assemblies showed many of the D4Z4 arrays to be incomplete or split across multiple scaffolds (Supplementary Table S2), creating difficulties for accurate D4Z4 counting and haplotyping. Thus, based on evidence from the raw reads, we find different D4Z4 repeat numbers to those reported previously (49). Additionally, we used the pipeline to process the publicly-available raw Nanopore data used for the CHM13v2.0 assembly (47), and found the number of repeat units in CHM13v2.0’s assembly of the 10q array (33 full 3.3 kb units) to be inconsistent with the raw Nanopore reads (32 full 3.3 kb units). This shows that there are still outstanding issues in the automated assembly of macrosatellite repeat regions, and demonstrates the need for a tool to visualise the repeat structure of raw reads.

We then used the pipeline to analyse Nanopore ultra-long whole-genome sequencing (WGS) data from the fibroblasts of two healthy controls, three patients diagnosed with FSHD1, three patients diagnosed with FSHD2, and two patients diagnosed with BAMS (Table 1). The D4Z4 haplotypes of the FSHD1 and FSHD2 patients have previously been studied using Southern blotting and/or molecular combing (50–54), with one of the FSHD1 patients exhibiting somatic mosaicism for a pathogenic 4qA 2RU allele (25% based on Southern blot) and another of the FSHD1 samples harbouring a 10qA *cis* triplication allele (53) (Supplementary Table S3). The D4Z4 haplotypes of the controls and BAMS patients have not previously been studied.

**Table 1.** Haplotyping results and number of spanning reads for D4Z4 alleles from FSHD, BAMS and control fibroblasts, from whole-genome and Cas9-targeted Nanopore sequencing.

| SAMPLE | 4q |  |  |  |  | 10q |  |  |  |
| --- | --- | --- | --- | --- | --- | --- | --- | --- | --- |
|  | Haplotype | No. RUs <sup>a</sup> | No. of WGS spanning reads | No. of Cas9 spanning reads | Upstream D4S2463 <sup>b</sup> | Haplotype | No. RUs | No. of WGS spanning reads | No. of Cas9 spanning reads |
| **Control** |  |  |  |  |  |  |  |  |  |
| AG09309 | 4qB | ≥38 | 0 | -- <sup>c</sup> | N | 10qA | 10 | 2 | -- |
|  | N/A <sup>d</sup> | N/A | 0 | -- | N/A | 10qA | 21 | 1 | -- |
| AG10803 | 4qB | 18 | 2 | -- | Y | 10qA | 10 | 2 | -- |
|  | 4qB | ≥35 | 0 | -- | N | 10qA | 13 | 2 | -- |
| **FSHD1** |  |  |  |  |  |  |  |  |  |
| 12566 | 4qAS | 3 | 4 | 20 | Y | 10qA | 9 | 4 | 12 |
|  | 4qAS | 42 | 1 | 0 | N | 10qB | 19 | 2 | 0 |
| FSHD1_3 | 4qAL | 1 | 3 | -- | Y | 10qA | 15+2+5 <sup>e</sup> | 1+1+2 <sup>f</sup> | -- |
|  | 4qB | 17 | 2 | -- | Y | 10qA | 35 | 1 | -- |
| 17706 | 4qAL | 1 | 0 | 53 | N | 10qA | 6 | 4 | 139 |
|  | 4qAL | 15 | 1 | 80 | Y | 10qA | 14 | 6 | 115 |
|  | 4qAL | 40 | 0 | 12 | N |  |  |  |  |
| 19187 | 4qAS | 2 | -- | 118 | -- | 10qA | 15 | -- | 33 |
|  | 4qAS | 22 | -- | 6 | -- | 10qA | 26+1 <sup>e</sup> | -- | 12 |
|  | 4qAM | 32 | -- | 8 | -- |  |  |  |  |
| **FSHD2** |  |  |  |  |  |  |  |  |  |
| 15166 | 4qAS | 12 | 4 | -- | N | 10qB | 13 | 2 | -- |
|  | 4qB | 21 | 2 | -- | N | N/A | N/A | 0 | -- |
| 34140 <sup>g</sup> | 4qAS | 25 | 1 | -- | Y | 10qB | 17 | 3 | -- |
|  | 4qAL | ≥20 | 0 | -- | N | 10qA | 19 | 3 | -- |
| 11491 | 4qAL | 22 | 1 | -- | N | 10qA | 6 | 1 | -- |
|  | N/A | N/A | 0 | -- | N/A | 10qA | 7 | 1 | -- |
| 11440 | 4qAS | 11 | -- | 14 | -- | 10qA | 14 | -- | 3 |
|  | 4qAS | 34 | -- | 4 | -- | 10qA | 19 | -- | 4 |
| **BAMS** |  |  |  |  |  |  |  |  |  |
| BAMS-1 | 4qAM | 32 | 0 | -- | N | 10qA | 12 | 0 | -- |
|  | 4qAS | 47 | 0 | -- | N | 10qA | 23 | 3 | -- |
| BAMS-9 | 4qB | 13 | 5 | 100 | Y | 10qA | 7 | 1 | 192 |
|  | 4qB | 26 | 2 | 27 | Y | 10qA | 31 | 1 | 9 |

<sup>a</sup> Refers to the number of complete 3.3kb KpnI-KpnI D4Z4 units (excluding partial D4Z4 units at the proximal and distal ends)
<sup>b</sup> Refers to whether D4S2463, an inverted and truncated D4Z4 unit found ~42kb upstream of 4q D4Z4 arrays, is captured in spanning reads from the WGS data
<sup>c</sup> Dashes (--) indicate that no WGS/Cas9-targeted sequencing was performed for that sample
<sup>d</sup> 'N/A' indicates that sequencing was performed, but the allele was not captured
<sup>e</sup> Alleles with in-cis duplications/triplications of the D4Z4 array
<sup>f</sup> Number of reads for the triplication allele spanning 1, 2 and all 3 of the arrays respectively
<sup>g</sup> Previously diagnosed as FSHD2 based on clinical findings and absence of a contracted D4Z4 array

The pipeline was able to successfully highlight and haplotype 4q and 10q reads from the patient fibroblasts (Table 1), with results having variable concordance with those from Southern blot and molecular combing (Supplementary Table S3). WGS captured spanning reads for 30 out of 41 alleles, with a maximum of six spanning reads being obtained for a single allele. We captured spanning reads corresponding to the pathogenic, contracted allele for two of the FSHD1 samples, but not for the mosaic FSHD1 sample, perhaps due to the contracted allele having a lower mosaic proportion (25%) (Figure 2A). The interactive visualisation tool also enabled assessment of the length of the distal D4Z4 unit, and we detected both the previously-reported 4qAS (~0.3 kb) and 4qAL (~1.9 kb) haplotypes. We additionally identified a new, previously unreported 4qA haplotype with a distal D4Z4 unit of length ~0.6 kb, present in the BAMS1 sample, which we designate ‘4qAM’ (Figure 2B). All 10qA alleles contained a distal D4Z4 unit of length ~0.3 kb, similar to 4qAS alleles.

Ultra-long read sequencing also enabled assessment of extended regions of 4q and 10q sequence upstream and downstream of the D4Z4 array (Figure 2B). For eight of the 4q alleles, reads were obtained that captured the inverted D4Z4 unit (D4S2463) containing *DUX4c* that is found ~42 kb upstream of the 4q D4Z4 array (55, 56) (Table 1, Figure 2B). For three of the samples (12566, 15166, 34140), 10q alleles were identified that belonged to B haplotypes and contained XapI restriction sites, indicating possible 4q-to-10q translocations upstream of the array. The region of homology between 4q and 10q extends ~42 kb upstream of the D4Z4 array (53, 57), and these alleles were able to be confidently assigned to chromosome 10q as the ultra-long reads spanned >42 kb of upstream sequence (Figure 2B). Moreover, the pipeline identified reads that spanned all three arrays of the 10qA in-*cis* triplication allele of sample FSHD1_3 (37-I_1_ in (53)), revealing the allele’s complex structure with nucleotide resolution (Figure 2B). As has been previously suggested, translocated or duplicated alleles with intact *DUX4* and PAS sequences may give rise to *DUX4* expression (16), however these complex alleles cannot always be accurately resolved by Southern blotting and molecular combing (15, 53). This demonstrates that our pipeline can comprehensively characterise 4q and 10q D4Z4 alleles, including reliable identification of contracted 4qA alleles to confirm the diagnosis of FSHD1, and identification of other structural variants which may contribute to FSHD pathogenesis.

### Cas9-targeted sequencing of 4q/10q D4Z4 alleles and FSHD2-associated genes

While ultra-long WGS is able to capture reads that span the entire 4q and 10q D4Z4 arrays with no prior assumption of the repeat composition or structure, a single PromethION flow cell per patient only provides low coverage. This can lead to pathogenic alleles being missed (Table 1, Figure 2A), difficulty in creating an accurate consensus sequence of each allele, and in exploring cell-to-cell epigenetic variability or indeed genetic mosaicism.

To address this, we used Cas9-targeted sequencing to enrich for the 4q and 10q D4Z4 regions, aiming to capture the whole set of alleles in each patient. We designed guide RNAs targeting regions upstream and downstream of the D4Z4 array, for both A and B alleles (Supplementary Table S4). Additionally, we complemented this with guide RNAs designed to target a panel of FSHD2-associated genes (*SMCHD1, DNMT3B* and *LRIF1*) (Supplementary Table S4), with a view to developing an all-in-one genetic and epigenetic research and diagnostic tool for FSHD. This enabled us to obtain high-read-depth sequencing of both chr 4q35 and 10q26 (Table 1, Figure 2A) and of *SMCHD1, DNMT3B* and *LRIF1* exons, respectively (Supplementary Figure S1).

For all except one of the samples assayed using Cas9-targeted sequencing (three FSHD1 samples, one FSHD2 sample, and one BAMS sample), we were able to determine the full set of 4q and 10q haplotypes by capturing at least one spanning read from each of the patient’s 4q/10q alleles (Table 1). This included the capture of all three 4q alleles from the two mosaic FSHD1 patients (Table 1, Figure 2A). While the proportions of spanning 4q reads for each allele from the mosaic samples did not match the expected mosaic proportions, sample 19187’s 4qA 2RU allele was captured at a much higher frequency than sample 17706’s 4qA 1RU allele, perhaps reflecting its higher mosaic proportion (50% vs 25%). Overall, there were far more spanning reads for shorter arrays than for longer arrays, likely due to the greater potential for fragmentation of long reads during library preparation, and preference for short reads during sequencing. Nevertheless, we were able to capture multiple spanning reads for alleles of up to 40 repeat units (~142 kb), well beyond the maximum read length for Cas9-targeted sequencing recommended by Oxford Nanopore Technologies (30 kb).

### Genetic analysis of 4q/10q alleles using high-accuracy consensus sequences

For alleles with high read depth from Cas9-targeted sequencing, we used Racon (25) to create full-length consensus sequences for the D4Z4 array. While many automated assembly tools struggle to accurately reconstruct repetitive alleles (Supplementary Table S2), our consensus-calling approach (see Methods) was able to correct errors while faithfully retaining the structure of the raw D4Z4 reads, including the number of repeat units and unusual variants such as a truncated internal D4Z4 repeat (Supplementary Figure S3C). This was able to be readily verified by using our interactive visualisation tool (Figure 2) to compare the repeat structure of the consensus sequences to that of the raw reads.

Using the consensus sequences, we were able to reliably analyse single-nucleotide variants (SNVs) and small insertions/deletions (indels) from 4qA, 4qB, and 10qA alleles. We identified 4q- and 10q-specific variants that were consistent with those seen in CHM13v2.0, including the two SNVs responsible for the 4q-specific XapI site and 10q-specific BlnI site (Figure 3A). Additionally, we identified qB-specific variants from the BAMS9 4qB alleles, that were consistent with those found in the 4qB allele from GRCh38 (Figure 3A). Several haplotype-specific variants were present in the *DUX4* open reading frame (ORF), including four silent SNVs, a 4qB-specific A>G variant leading to an I229V substitution, and a 10qA-specific 6-bp deletion leading to A340_S341del. Several variants were also identified in the *DUX4* promoter region, but none were found to overlap known promoter elements including the GC box, TACAA box, and initiator sequence (Supplementary Figure S2).

**Fig. 3.**
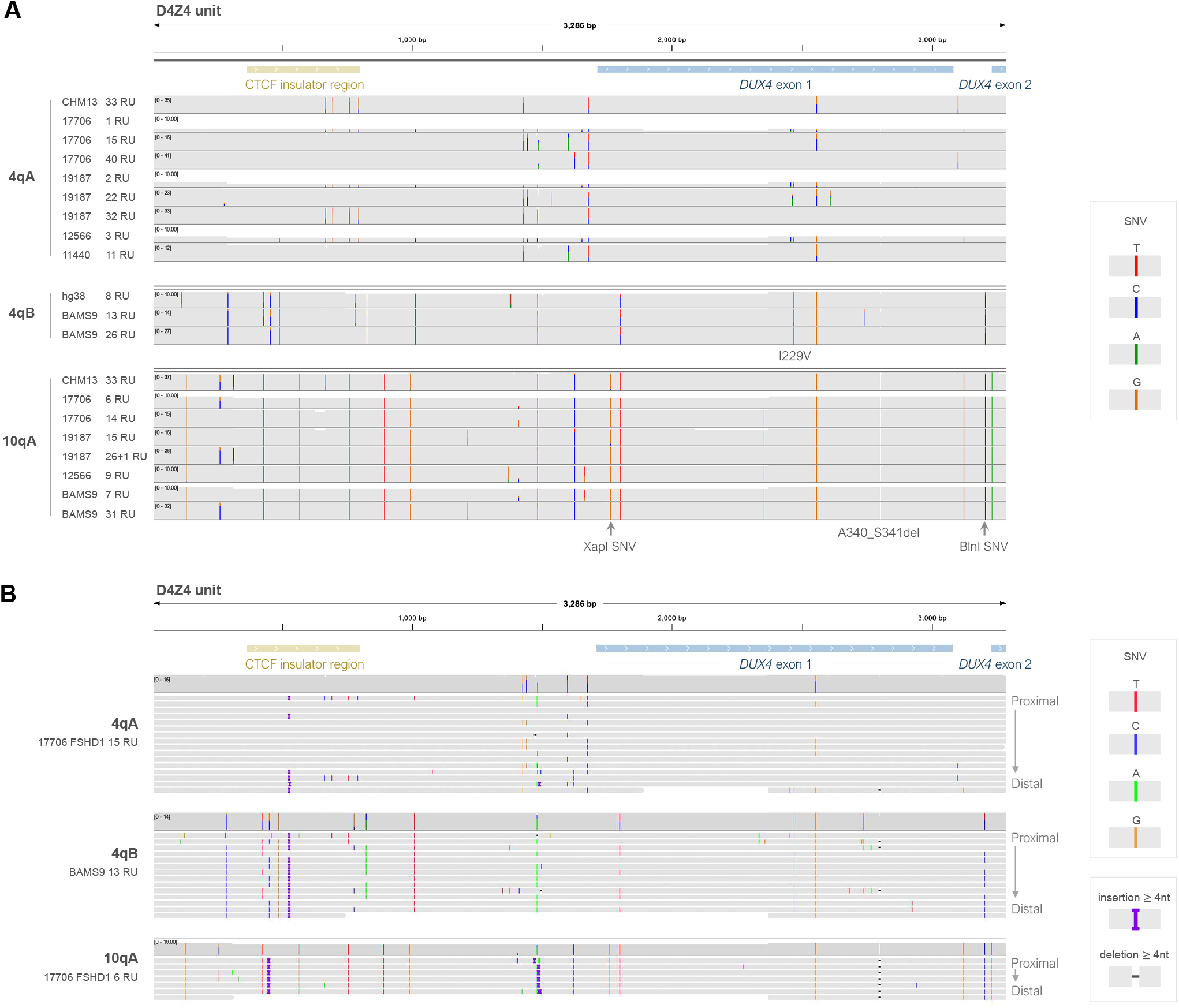
Array-wide and base-level analysis of D4Z4 genetic variants using consensus sequences from high-coverage long-read sequencing. (A) IGV coverage plots for alignments of D4Z4 units from the consensus sequences for patient 4q and 10q alleles, showing 4qA-, 4qB- and 10qA-specific genetic variants consistent with CHM13v2.0 and GRCh38. (B) IGV alignments of D4Z4 units from the consensus sequences for representative 4qA, 4qB and 10qA alleles from samples 17706 and BAMS3. Within each alignment track, full-length D4Z4 units are arranged from top to bottom in order of proximal to distal position within the array. Indels <4nt which were not consistent between repeat units were hidden to aid in visualisation (Supplementary Figure S3). Consensus sequences were generated using Racon using spanning reads from high-coverage Cas9-targeted Nanopore sequencing. SNV: single nucleotide variant.

Comparison of the variants between individual D4Z4 units within each 4q and 10q array showed 4q arrays to be more heterogeneous than 10q arrays (Figure 3B, Supplementary Figure S3), consistent with previous observations (49). For both 4qA and 4qB arrays, the proximal repeat unit contained a distinct set of variants compared to most other D4Z4 units in the array, although these variants were occasionally also found in internal repeats (Figure 3B). Variants in the proximal and internal repeat units have previously been used to define D4Z4 subhaplotypes (21), however these have largely been restricted to SNVs affecting restriction enzyme sites. The additional 4qA-, 4qB- and 10qA-specific genetic variants identified here provide further insight into D4Z4 subhaplotypes and the composition of the D4Z4 array.

The composition of the D4Z4 array could also be used to reconstruct full-length haplotypes for alleles without spanning reads, and we demonstrated this for the BAMS1 4qAM allele (Supplementary Figure S4). Interestingly, we also identified a 4qAM allele from the Cas9-sequencing data for sample 19187 (FSHD1) (Figure 4), and we found the full-length BAMS1 4qAM allele to be the same as 19187’s 4qAM allele (Supplementary Figure S3C), suggesting a common ancestral origin.

**Fig. 4.**
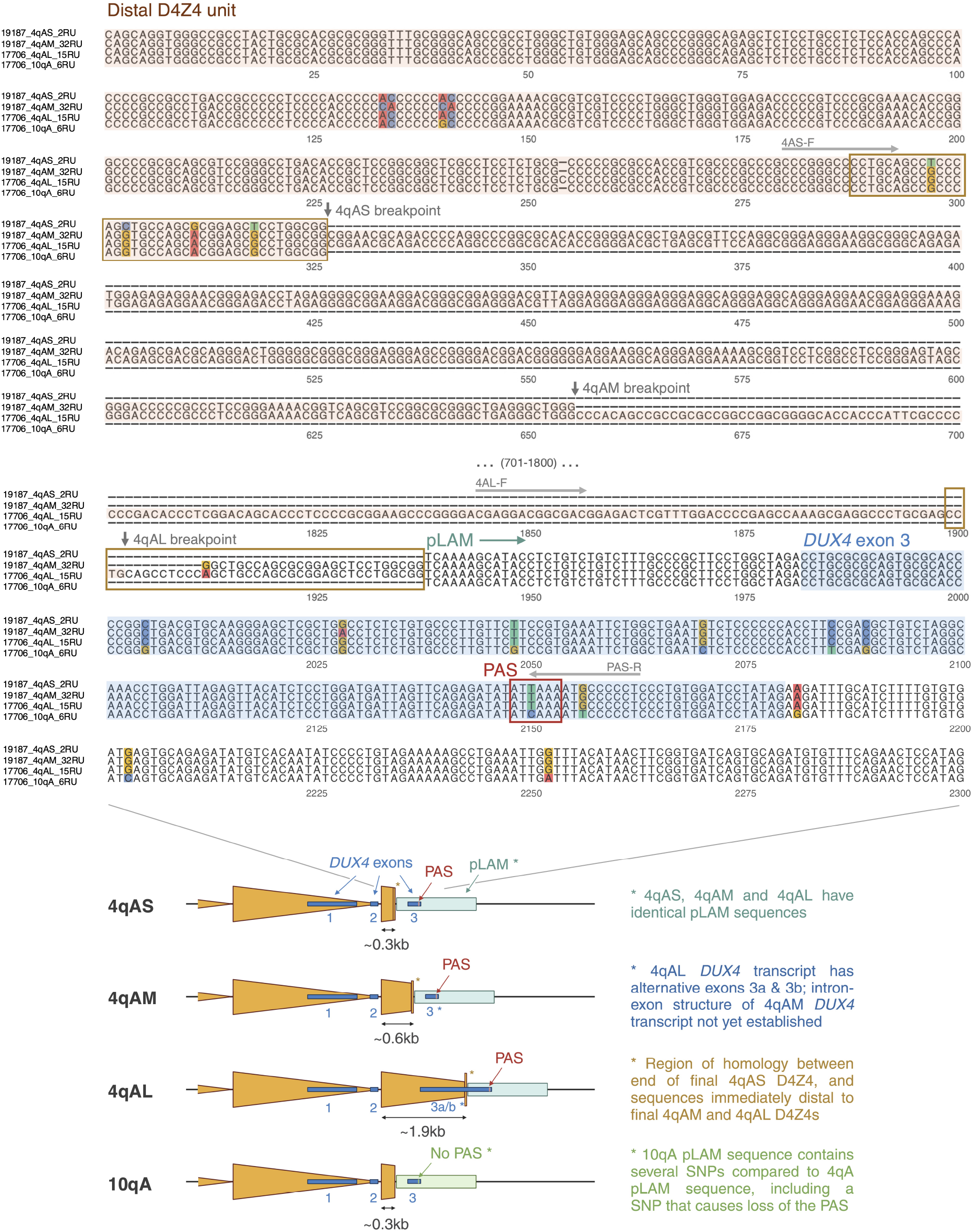
Base-level comparison of distal D4Z4 sequences from 4qAS, 4qAM, 4qAL and 10qA alleles. (A) Alignment of distal D4Z4 sequences beginning from the proximal KpnI site for the final partial D4Z4 unit, and extending into the proximal portion of the pLAM sequence containing DUX4 exon 3. Breakpoints for the final partial D4Z4 units (vertical arrows) were determined by alignment to a full-length D4Z4 sequence. The start of pLAM is marked as the first nucleotide following the final partial 4qAS D4Z4 unit. The 4qAM and 4qAL alleles contain an extra length of sequence between the end of their final partial D4Z4 units and the start of pLAM, which corresponds to the end of the 4qAS allele’s final partial D4Z4 unit (shown in orange boxes). Locations of 4qAS- and 4qAL-specific forward PCR primers (4AS-F, 4qAL-F) and a polyadenylation signal (PAS) reverse primer (PAS-R) used in (22) are shown above their respective sequences. Sequences are extracted from consensus sequences generated using Racon from high-coverage Cas9-targeted Nanopore sequencing.

The consensus sequences also enabled more detailed assessment of the 4q subhaplotype based on sequences proximal and distal to the D4Z4 array. Alignment of the distal D4Z4 sequences of 4qAS, 4qAL and 4qAM alleles showed all three haplotypes to have identical pLAM sequences, containing an intact PAS (Figure 4). The different length of the final partial repeat unit raises the possibility that 4qAM alleles give rise to different DUX4 transcripts. To further assess the potential pathogenicity of 4qAM alleles, we also assessed the simple sequence length polymorphism (SSLP) found ~3.5 kb upstream of the D4Z4 array that has been used to further delineate permissive and non-permissive D4Z4 subhaplotypes (58). We found that the SSLP for the BAMS1 4qAM allele corresponded to the 4A161 subhaplotype, which is permissive for FSHD. This suggests that 4qAM alleles may have the potential to be pathogenic, and may need to be considered in genetic testing.

These findings demonstrate the utility of our approach for analysing D4Z4 alleles at array-wide, base-level resolution, including the potential to further characterise known D4Z4 subhaplotypes, discover new subhaplotypes, and resolve the composition of D4Z4 arrays.

### Detection of pathogenic variants in *SMCHD1, LRIF1* and *DNMT3B*

We next applied a variant calling and variant effect prediction pipeline to the whole-genome and Cas9-targeted Nanopore sequencing data, to assess whether our assay could obviate the need for separate genetic testing. We focused on a panel of FSHD2-associated genes, *SMCHD1, LRIF1* and *DNMT3B*, part of the gene panel for Cas9-mediated enrichment. *SMCHD1* genotyping has previously been performed for the patients included in this study (Supplementary Table S3).

We identified pathogenic *SMCHD1* variants for three of the four FSHD2 samples (11440, 15166, 11491) and both of the BAMS samples (BAMS1, BAMS9) that recapitulated the prior genotyping results (Supplementary Table S3). For one of the patients diagnosed as FSHD2 (34140), no pathogenic variant in *SMCHD1, DNMT3B* or *LRIF1* was found. The presence of a synonymous variant was confirmed (p.L1031), but with no further evidence of pathogenicity, calling into question the diagnosis as FSHD2.

The BAMS variants were heterozygous missense variants located in or adjacent to *SMCHD1*’s N-terminal AT-Pase domain (sample BAMS1: p.E136G, sample BAMS9: p.D420V). Conversely, the FSHD2 variants were heterozygous splice donor (sample 11440: c.2338+4A>G (intron 18); sample 11491: c.5547+5G>A (intron 44)) or frameshift (sample 15166: c.4608_4614dup, p.Ala1539Tyrfs*6 (exon 37)) variants found closer towards the C-terminal end of the protein, predicted to lead to *SMCHD1* haploinsufficiency and consistent with an FSHD2 diagnosis.

In one of the FSHD1 samples (12566), we also identified a previously unreported p.R1887L missense variant in exon 45 of *SMCHD1* which was predicted to be pathogenic by AlphaMissense (59), with a pathogenicity score of 0.9642. None of the samples had pathogenic variants in *LRIF1* or *DNMT3B*. Notably, whereas some FSHD2-associated genes had only low coverage with whole-genome sequencing, the read depth was significantly increased with Cas9-targeted sequencing, aiding in detection of heterozygous pathogenic variants.

This confirmed the suitability of a single nanopore sequencing assay for 4q haplotyping, 4q structural characterisation and variant detection in an FSHD gene panel.

### Allele-specific CpG methylation patterns across the D4Z4 array

#### Array-size-dependent methylation patterns are seen in FSHD1 and control fibroblasts

To assess the methylation patterns of D4Z4 arrays of different lengths and *SMCHD1* variant status, we called CpG methylation for the 4q and 10q reads, and used NanoMethViz (26, 27) to plot the methylation of individual molecules as well as the smoothed methylation profile across the complete array for each allele.

For FSHD1 and control samples, an oscillatory pattern of D4Z4 methylation was seen within each repeat unit, with low levels of methylation around the DR1 region, which corresponds to the distal portion of a proposed CTCF binding site (37, 60), and high levels of methylation around the *DUX4* transcription start site (TSS) (Figure 5, Supplementary Figure S6), consistent with previous studies (40, 61). These oscillatory patterns were present in all non-contracted alleles, but absent from the hypomethylated, contracted FSHD1 alleles. Autocorrelation of the methylation status of CpGs across the D4Z4 array indicated strong correlation between sites separated by ~3.3 kbp, corresponding to the size of one D4Z4 unit, as well as between sites separated by ~180 bp (Supplementary Figure S7), which likely represents nucleosome positioning (40, 62). The high read depth provided by Cas9-targeted sequencing enabled us to see that for each allele, the overall methylation patterns were highly consistent between individual molecules (Figure 5), but that there was significant intramolecular heterogeneity at the level of individual CpG sites (Supplementary Figure S8).

**Fig. 5.**
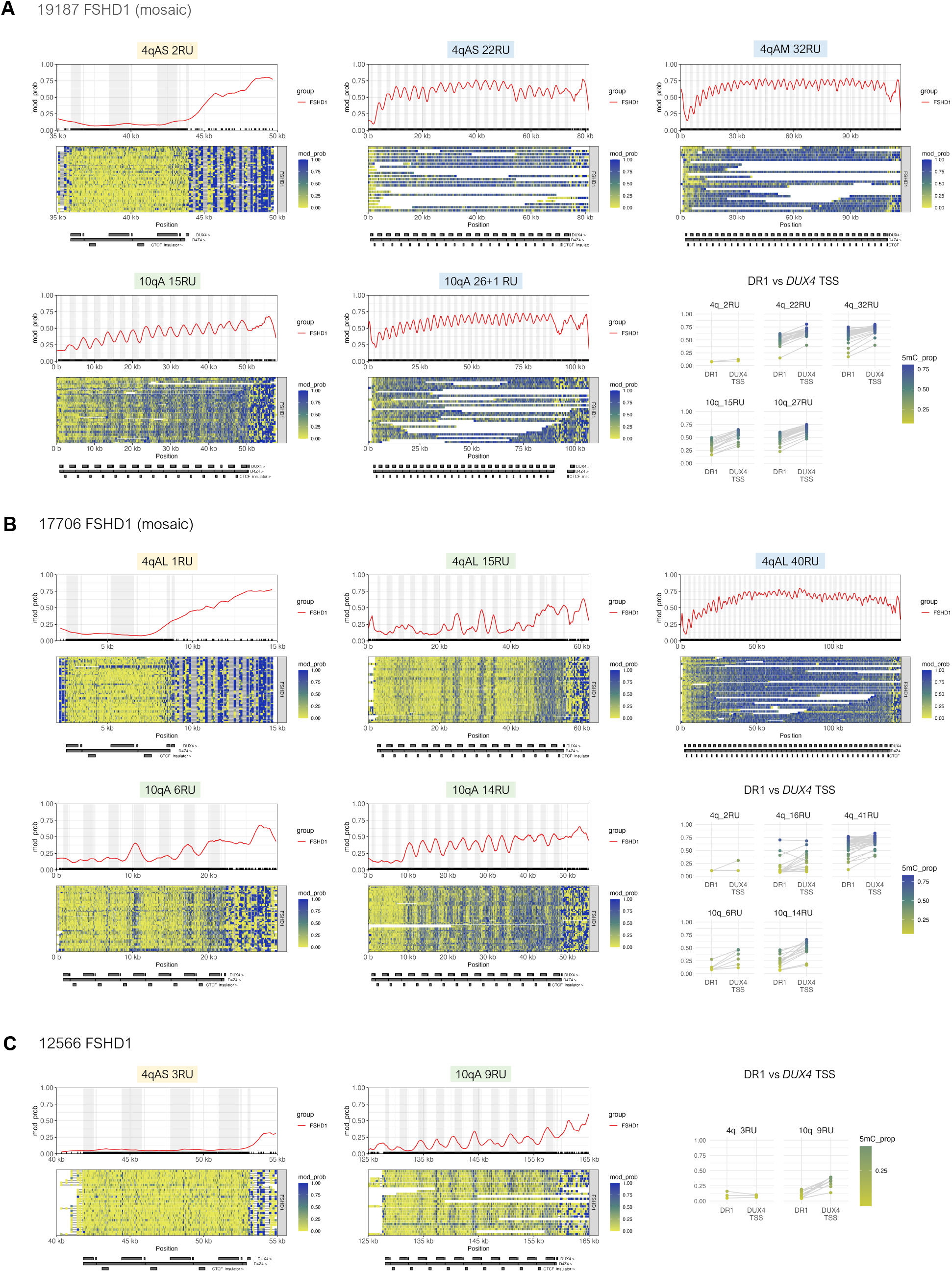
Allele-specific, array-wide D4Z4 methylation profiles for FSHD1 fibroblasts from Cas9-targeted sequencing. Single-molecule and smoothed methylation plots for 4q and 10q D4Z4 alleles from the FSHD1 samples (A) 19187, (B) 17706 and (C) 12566, generated using NanoMethViz (26, 27). The locations of *DUX4* exons are shaded in gray on the smoothed methylation plot. Annotations for D4Z4 repeat units, CTCF insulator regions, and *DUX4* exons are shown below each plot. Mean methylation rates over the DR1 region (positions 563–814 of the D4Z4 KpnI-KpnI unit (37) and *DUX4* transcription start site (TSS) region (−200 to +200 with respect to the TSS at position 1688 of the D4Z4 KpnI-KpnI unit) within each D4Z4 unit were also plotted for each allele. Points for DR1 and *DUX4* TSS regions from the same D4Z4 unit are connected by gray lines.

Differing array-wide methylation patterns were observed for short, intermediate and long D4Z4 arrays (Figure 5, Supplementary Figure S6). The short, pathogenic FSHD1 arrays, which ranged from 1-3 repeat units, were severely hypomethylated across the full length of the array (Figure 5 and Supplementary Figure S6, alleles shaded in yellow). Long arrays of >20 repeat units were much more highly methylated, and tended to display a regular, stepwise increase in D4Z4 methylation over the first ten or so repeat units before reaching a constant, high level of methylation (Figure 5 and Supplementary Figure S6, alleles shaded in blue), similarly to what has been described previously (40). By contrast, short D4Z4 arrays at the upper end of the pathogenic range for FSHD1 (6-10 repeat units) and intermediate-length arrays (11-20 repeat units) were more variable in their methylation patterns (Figure 5 and Supplementary Figure S6, alleles shaded in green). While these arrays were often more highly-methylated at their distal ends compared to their proximal ends, this proximal-to-distal increase in methylation did not always occur in a stepwise fashion, as exemplified by the ‘irregular’ methylation patterns seen in several of the alleles from sample 17706 (Figure 5B). These observations may partially explain why array sizes in the 1-4 repeat unit range are seen in more clinically-severe and early-onset cases of FSHD, while longer arrays are associated with variable penetrance and a broad range of clinical severity (29, 64).

The differences in D4Z4 methylation between short, intermediate and long D4Z4 arrays were also reflected in the global D4Z4 methylation level for each allele, as measured by the proportion of all methylated CpGs across the D4Z4 array (henceforth ‘global D4Z4 5mC rate’). Overall, we found a positive correlation between the global D4Z4 5mC rate and the number of repeat units for both FSHD1 samples and controls (Pearson correlation coefficient 0.844, Spearman correlation coefficient 0.826) (Figure 6A). As only the terminal *DUX4* gives rise to stable transcripts in permissive 4qA alleles, we additionally assessed the mean methylation of the final full D4Z4 unit, which contains the promoter and exons 1 and 2 of the terminal *DUX4*. Amongst FSHD1 and control samples, there was a moderate correlation between the number of repeat units and mean methylation rate of the final full D4Z4 unit (‘final D4Z4 5mC rate’) (Pearson correlation coefficient 0.701, Spearman correlation coefficient 0.732) (Figure 6B). As expected, the pathogenic, contracted FSHD1 alleles had very low final D4Z4 5mC rates, ranging from 0.064 to 0.097. However, there were two intermediate-length *SMCHD1*-wildtype alleles which also had low final D4Z4 5mC rates, the FSHD1_3 4qB 17RU allele (final D4Z4 5mC rate 0.118) and the AG10803 (ag10) 4qB 18RU allele (final D4Z4 5mC rate 0.149). Interestingly, the AG10803 4qB 18RU allele appeared to be hypomethylated across the full length of the array (global D4Z4 5mC rate 0.116), although there were only two spanning reads to support the methylation signal (Supplementary Figure S6A). These findings once again suggest that in FSHD1 and control fibroblasts, while contracted D4Z4 arrays are consistently hypomethylated, and long D4Z4 arrays are consistently hypermethylated, methylation patterns for intermediate-length arrays are more variable.

**Fig. 6.**
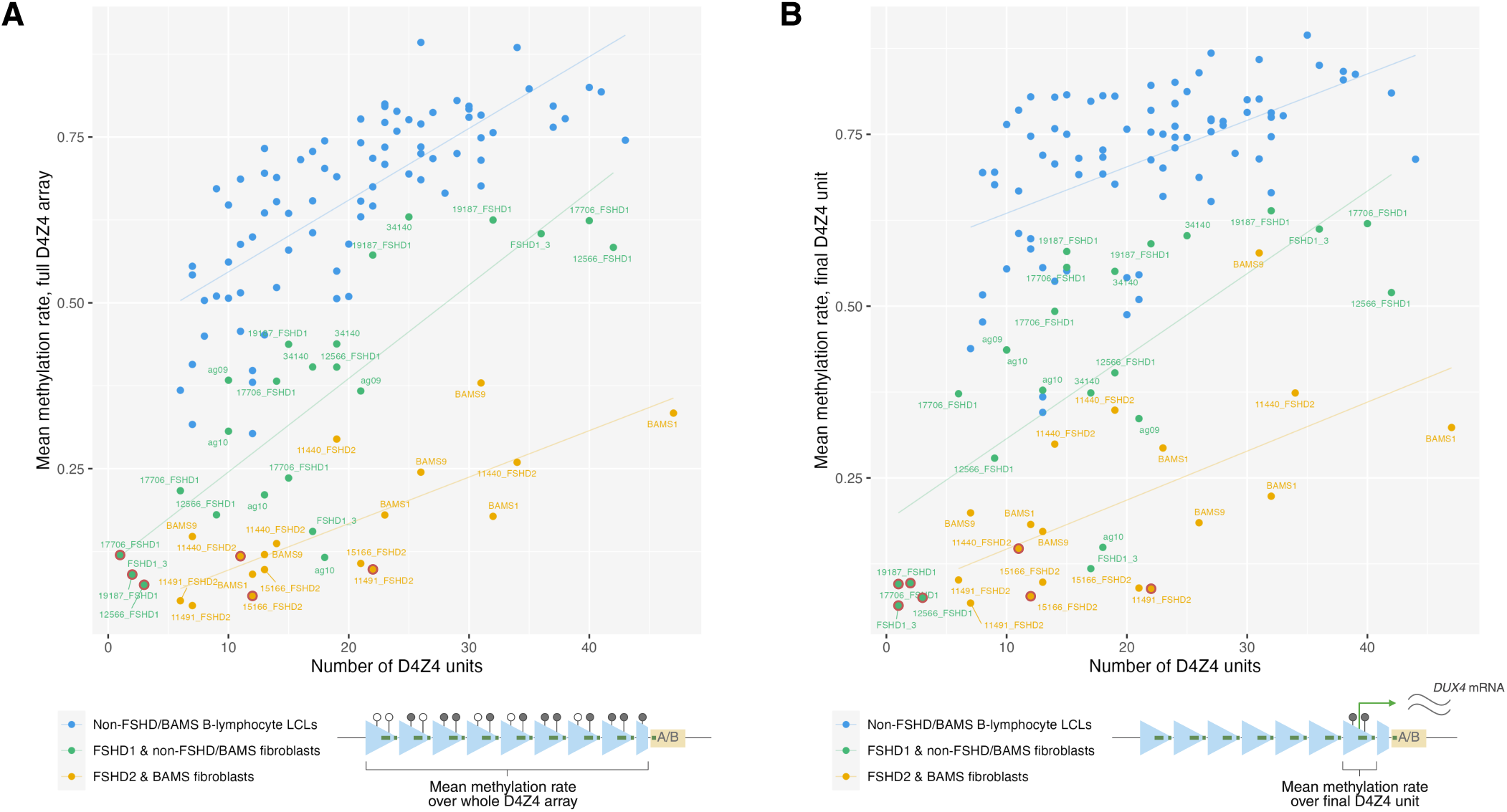
Correlation of overall D4Z4 methylation levels with the number of repeat units. Mean methylation rates across (A) the full 4q/10q D4Z4 array or (B) the final D4Z4 unit against the number of D4Z4 units, plotted for alleles from *SMCHD1*-wildtype fibroblasts (FSHD1 and non-FSHD/BAMS), pathogenic-*SMCHD1*-variant fibroblasts (FSHD2 and BAMS), and 30 B-lymphocyte lymphoblastoid cell lines (LCLs) from the 1000 Genomes Project (63). Sample 34140, initially clinically diagnosed as FSHD2, was included in the ‘non-FSHD/BAMS’ group based on the lack of pathogenic *SMCHD1, LRIF1* or *DNMT3B* variants and overall methylation profile. Mean methylation rate was calculated as the total number of methylated CpGs from all reads / the total number of methylated + unmethylated CpGs from all reads, across the defined regions. Regression lines were plotted for each group of samples. Alleles presumed to be the pathogenic alleles for each of the FSHD patients are indicated with red rings. (A) Pearson and Spearman correlation coefficients for FSHD1 and non-FSHD/BAMS fibroblasts, FSHD2 and BAMS fibroblasts, and B-lymphocyte LCLs were 0.844 and 0.826, 0.757 and 0.742, and 0.746 and 0.798, respectively. (B) Pearson and Spearman correlation coefficients for FSHD1 and non-FSHD fibroblasts, FSHD2 and BAMS fibroblasts, and B-lymphocyte LCLs were 0.701 and 0.732, 0.583 and 0.607, and 0.531 and 0.552, respectively.

### B-lymphocyte lymphoblastoid cell lines show similar D4Z4 methylation patterns to FSHD1 and control fibroblasts, but have higher methylation levels

To assess whether these length-dependent methylation patterns are also seen in a larger cohort and other cell types, we analysed D4Z4 methylation in 30 B-lymphocyte lymphoblastoid cell lines (LCLs) from the 1000 Genomes Project (1KGP), using publicly-available Nanopore WGS data from the 1KGP ONT Sequencing Consortium (1KGP-ONT) (63). We found 83 4q/10q alleles with reads that spanned the whole D4Z4 array, ranging in length from 6 to 43 repeat units (Supplementary Table S5). The patterns of DNA methylation across the array in the B-lymphocyte LCLs were similar to those seen in *SMCHD1*-wildtype fibroblasts. All arrays had oscillatory patterns of low methylation around the DR1/CTCF insulator region and high methylation around the *DUX4* TSS (Supplementary Figure S10). For long arrays (>20 repeats), methylation levels increased over the first ten or so repeat units then plateaued at a constant, high level across the remainder of the array, or were consistently high across the whole array. Short and intermediate-length arrays (6-20 repeat units) tended to display more regular, stepwise increases in methylation from the proximal to distal end of the array, contrasting with the ‘atypical’ methylation patterns seen in several of the alleles from the FSHD1 and control fibroblasts. Similarly to what we found in fibroblasts, there was a positive correlation between the global D4Z4 5mC and final D4Z4 5mC rates, and the number of D4Z4 units (Figure 6). However, mean D4Z4 methylation rates were generally higher in the B-lymphocyte LCLs compared to the *SMCHD1*-wildtype fibroblasts, suggesting that global D4Z4 methylation levels are cell-type specific.

#### Both FSHD2 and BAMS fibroblasts have global D4Z4 hypomethylation

We sought to investigate the effect of pathogenic FSHD2 and BAMS *SMCHD1* variants on D4Z4 methylation. In both FSHD2 and BAMS, there was global hypomethylation of D4Z4 arrays compared to similar-length FSHD1 and control samples (Figure 6A). Despite the differing effects of FSHD2 and BAMS variants on the *SMCHD1* protein (predicted haploinsufficiency and missense mutations in the ATPase domain, respectively), FSHD2 and BAMS samples showed similar overall methylation levels and methylation patterns to each other (Figure 7, Supplementary Figure S9). Short and intermediate-length arrays were generally severely hypomethylated across the full length of the array. Longer alleles had relatively higher methylation levels, yet were still markedly hypomethylated compared to similar-length *SMCHD1*-wildtype samples (Figure 6A), and generally had lower methylation rates for their final D4Z4 units (Figure 6B) owing to minimal proximal-to-distal increase in array-wide methylation. This was clearly demonstrated by comparison of the 32RU 4qAM alleles from BAMS9 and 19187_FSHD1 (Figure 5A, Figure 7B). However, interestingly, we also identified several reads with very high methylation levels that mapped to the BAMS1 47RU 4qAS allele (Supplementary Figure S11), suggesting either the presence of a separate, highly-methylated cell population, or an unexpected and sporadic ‘rescue’ of these cells from hypomethylation.

**Fig. 7.**
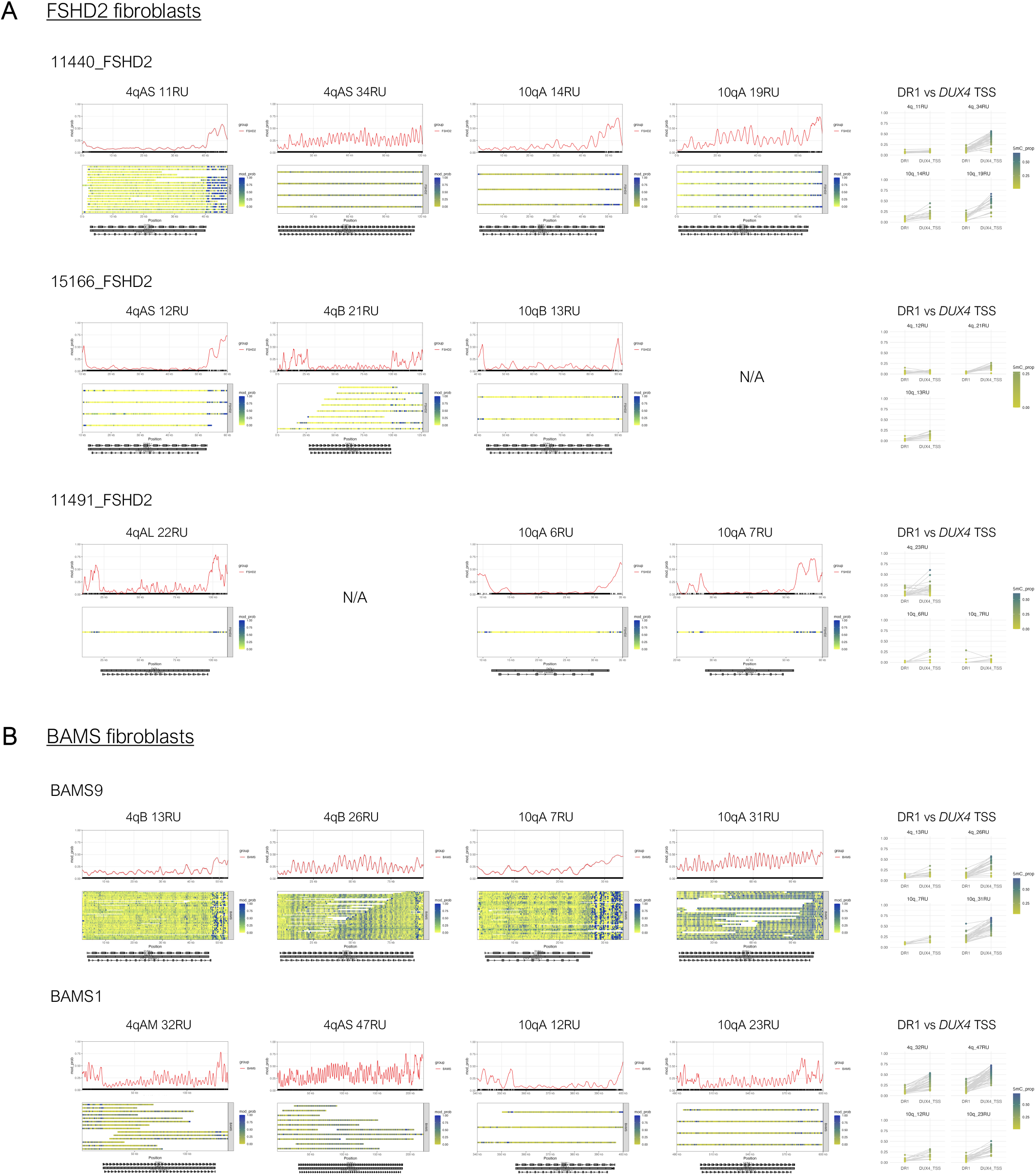
Allele-specific, array-wide D4Z4 methylation profiles for FSHD2 and BAMS fibroblasts. Single-molecule and smoothed methylation plots for 4q and 10q D4Z4 alleles from (A) FSHD2 and (B) BAMS samples, generated using NanoMethViz (26, 27). Annotations for D4Z4 repeat units, CTCF insulator regions, and *DUX4* exons are shown below each plot. Mean methylation rates over the DR1 region (positions 563–814 of the D4Z4 KpnI-KpnI unit (37) and *DUX4* transcription start site (TSS) region (−200 to +200 with respect to the TSS at position 1688 of the D4Z4 KpnI-KpnI unit) within each D4Z4 unit were also plotted for each allele. Points for DR1 and *DUX4* TSS regions from the same D4Z4 unit are connected by gray lines. The full-length reference sequences for the BAMS1 4qAM and 4qAS alleles were determined as described in Supplementary Figures S4 and S11.

In terms of intra- and inter-repeat methylation patterns, oscillations between low methylation around the DR1/CTCF insulator region and high methylation around the *DUX4* TSS were variably present in hypomethylated alleles, but were prominent in more highly methylated longer alleles (Figure 7). Autocorrelation of 5mC signals once again indicated methylation patterns of ~3.3 kbp and ~180 bp periodicity (Supplementary Figure S7), suggesting that the mechanisms that give rise to these patterns are retained in FSHD2 and BAMS.

### Complete genetic and epigenetic analysis of in-*cis* duplicated arrays and upstream inverted D4Z4 units

While several studies have used molecular combing to study the structure of in-*cis* duplication alleles (16, 34, 36, 45, 53), this technique is unable to reveal the underlying genetic sequence or epigenetic status, and many aspects of the genetics and epigenetics of these complex alleles have yet to be resolved. Therefore, we sought to use long-read sequencing to further investigate the composition and methylation of in-*cis* duplicated arrays.

Our pipeline identified in-*cis* duplications downstream of the main D4Z4 array in two of the FSHD1 alleles (19187 10qA, FSHD1_3 10qA). Sample 19187’s complex 10qA allele was composed of a 26RU array followed by a 1RU array, separated by a ~6.5 kb spacer sequence, with the proximal partial D4Z4 of the duplicated array beginning ~0.3 kb after the KpnI restriction site (Figure 8A). Sample FSHD1_3’s complex 10qA allele contained an in-*cis* triplication, comprising a 15RU array followed by a 2RU array and a 5RU array, separated by two spacer sequences of ~20 kb, with the proximal partial D4Z4s of the duplicated arrays both beginning ~1.5 kb after the KpnI restriction site (Figure 8B). Analysis of the genetic sequence revealed all of the arrays to be 10qA-type, as determined by the presence of BlnI sites and 10qA-type pLAM sequences lacking a functional PAS. Moreover, the spacer sequences and the sequences distal to the duplicated array all corresponded to the sub-telomeric sequence that is normally found immediately distal to the 10qA D4Z4 array.

**Fig. 8.**
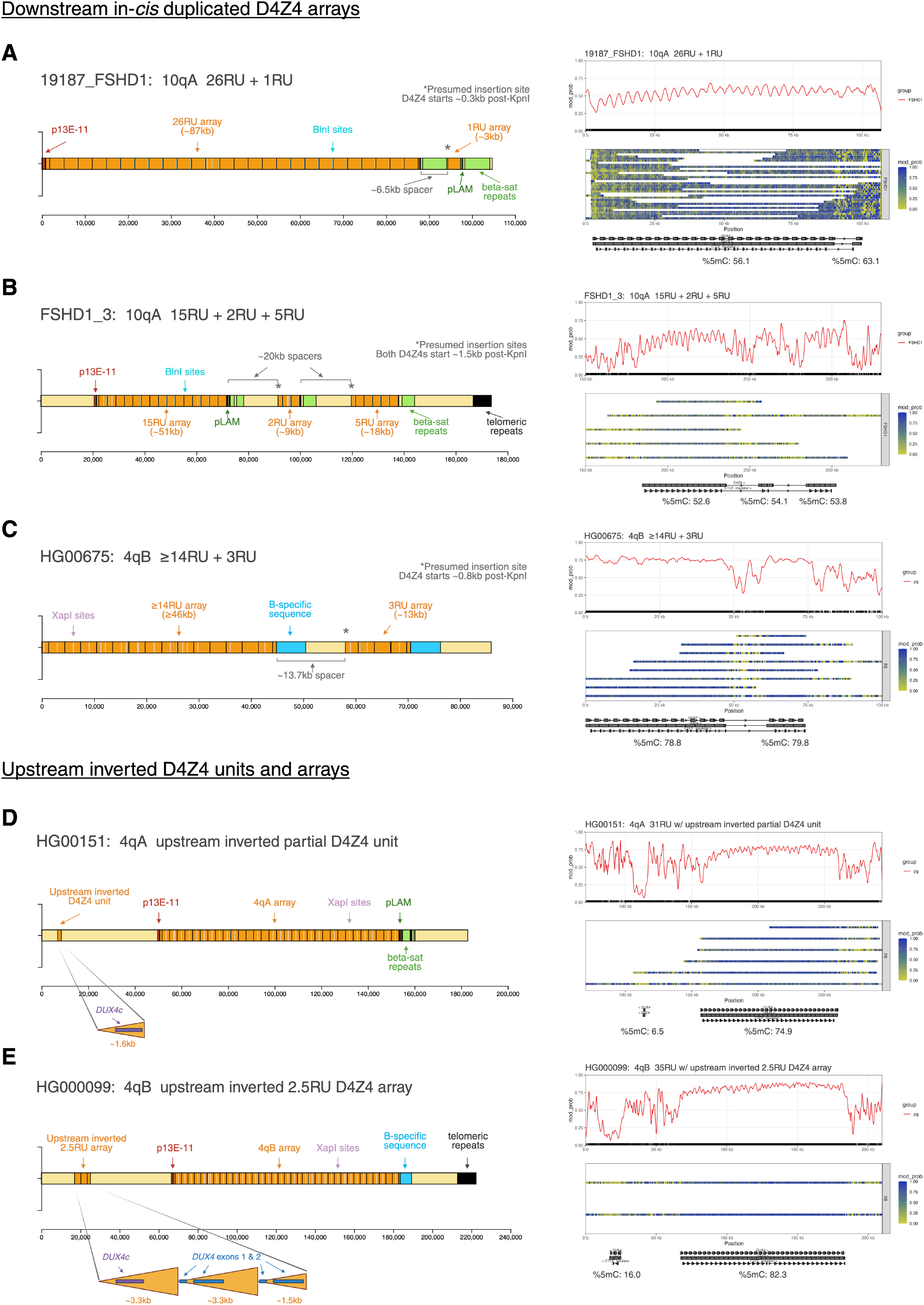
Analysis of the structure and methylation of duplicated D4Z4 arrays and upstream inverted D4Z4 units. (A), (B), (C) Structure of 4q and 10q alleles identified by the pipeline to have in-*cis* duplications downstream of the D4Z4 array, alongside smoothed methylation plots showing that these duplicated arrays are hypermethylated. The overall %5mC for each D4Z4 array is shown below the annotation track for the methylation plot. (D) Representative example of the inverted D4S2463 unit located *~*42 kb upstream of the 4q D4Z4 array, which is found to be hypomethylated in FSHD and control fibroblasts and B-lymphocyte LCL samples. (E) The HG00675 cell line from the 1000 Genomes Project (63) has a 4qB allele with an upstream inverted D4Z4 array of 2.5 repeat units in place of the D4S2463 unit. This upstream inverted array is also hypomethylated.

We also identified the first reported case of a 4qB in*cis* duplicated array, present in the B-lymphocyte LCL sample HG00675. The allele contained a proximal array of at least 14RU, followed by an in-*cis* duplicated array of 5RU. The arrays were separated by a ~13.7 kb spacer sequence, with the proximal partial D4Z4 of the duplicated array beginning ~0.8 kb after the KpnI restriction site (Figure 8C). Both arrays were 4qB-type, and once again, the spacer sequence was identical to the sequence distal to the duplicated array, both corresponding to the 4qB sub-telomeric sequence. These findings mirror those in Lemmers et al. (45), where the spacer sequence for 4qA-type in-*cis* duplication alleles was found to correspond to the 4qA sub-telomeric sequence. The shared basic structure of these 4qA, 4qB and 10qA duplication alleles suggests that they emerged via a common underlying mechanism, likely involving intrachromatid gene conversion.

We then assessed the methylation of the duplicated arrays. Interestingly, for all of these alleles, the downstream, contracted arrays were highly methylated, with average methylation levels similar to or higher than those of the non-contracted upstream arrays (Figure 8A-C). Additionally, for sample FSHD1_3’s in-*cis* triplication 10qA allele, the average methylation of the proximal 15RU array was higher than might be expected if it were an isolated 15RU array, based on comparison with the global D4Z4 5mC rate of sample FSHD1_3’s 4qB 17RU allele (0.155) and the methylation of intermediate-length arrays in other *SMCHD1*-wildtype samples (Figure 6A). Lemmers et al. (45) found a similar phenomenon for a mosaic 17+9RU 4qA allele and 17+2RU 4qA allele, where the proximal 17RU allele for the 17+9RU array was more highly-methylated than for the 17+2RU array. This may indicate the existence of bi-directional methylation spreading mechanisms between proximal and distal arrays, as suggested by Lemmers et al. (45), or reflect an underlying chromatin state that is dependent upon the additive effect of both duplicated and non-duplicated arrays.

To further characterise the structure and methylation of in-*cis* D4Z4 repeats, we also assessed the inverted D4Z4 unit (D4S2463) that is found ~42 kb upstream of the 4q D4Z4 array. Consistent with previous reports (55, 56), we found that D4S2463 is a truncated unit of ~1.5 kb that is homologous to the distal portion of the KpnI-KpnI D4Z4 unit, and contains a homologous *DUX4c* gene. Contrary to the downstream in-*cis* duplicated arrays, the upstream inverted repeat was consistently hypomethylated in the FSHD, BAMS and control fibroblast samples and in the B-lymphocyte LCL samples (Figure 8D).

Moreover, we identified one B-lymphocyte LCL allele (HG00099 4qB) that had an inverted D4Z4 array of 2.5 repeat units ~42 kb upstream of the main D4Z4 array, rather than a single D4S2463 unit (Figure 8E). Several cases of the same allelic variant have also previously been identified using molecular combing (16), where the inverted array was suggested to be an expansion of the D4S2463 repeat. Here, we were able to analyse the composition of the inverted array at base-level resolution, and we found that the first inverted D4Z4 unit was ~1.6 kb and corresponded to the distal ~1.6 kb of D4Z4, whereas the second and third inverted D4Z4 units were ~3.3 kb units containing *DUX4* and *DUX4c* sequences, respectively. Once again, Racon (25) was used to create a consensus sequence for the upstream inverted D4Z4 array, and it was found that all three repeat units contained 4qB-type SNVs, while the sequence for the last ~1.5 kb of the final repeat corresponded to the normal sequence for the inverted D4S2463 unit (Supplementary Figure S12). This suggests that the inverted array may have arisen by insertion of a segment of 4qB array at the breakpoint site for the D4S2463 unit, mediated by partial homology between the D4S2463 unit and the normal D4Z4 sequence; or alternatively, that the 4qB-type inverted array is in fact the ancestral allele, and gave rise to the partial D4S2463 repeat by contraction.

We then assessed the methylation of this inverted array. Similarly to the single inverted D4S2463 units, the inverted array was hypomethylated, with a mean 5mC rate of 0.160. Therefore, overall, we found that downstream, in-*cis* duplicated arrays were methylated at similar levels to the first D4Z4 array, whereas upstream inverted arrays were hypomethylated similar to the typical D4S2463 unit. This could suggest that methylation spreading mechanisms at the D4Z4 locus, if present, are orientation-dependent, or may provide further support for an insulator mechanism at the D4Z4 locus that separates upstream inverted and downstream non-inverted arrays into separate chromatin compartments (50, 60).

## Discussion

The D4Z4 macrosatellite array is one of the most difficult regions in the genome to study. Here we extend upon previous studies of D4Z4 using nanopore sequencing (40–46) to comprehensively characterise the array-wide genetic structure and methylation patterns of macrosatellite repeats, at base-level resolution. We have developed an optimised protocol and all-in-one analysis pipeline for FSHD, generating reads that span full-length D4Z4 arrays of up to 42 repeat units (~140 kb), as well as entire in-*cis* duplication and triplication alleles (~173 kb). Moreover, we designed guide RNAs (gRNAs) for Cas9-targeted sequencing that were able to enrich for both A- and B-type 4q and 10q arrays with high efficiency, identifying the full complement of patient 4q and 10q alleles, including cases of mosaicism, and to generate accurate consensus sequences for downstream analysis. Coupling this with gRNAs for FSHD2-associated genes (*SMCHD1, DNMT3B* and *LRIF1*) allowed us to accurately detect pathogenic FSHD2 gene variants in the same assay.

To aid in haplotyping and interpretation of the sequencing results, we developed a custom pipeline and interactive tool for annotation and visualisation of raw D4Z4 reads, outlining issues with current automated macrosatellite assembly. By annotating and exploring individual raw reads, we were able to more reliably resolve the structure, repeat number, and haplotype of D4Z4 alleles, which is critical for accurate downstream analyses and accurate FSHD diagnoses.

We demonstrated the efficacy of our pipeline on samples from several patients diagnosed with FSHD1, FSHD2 and BAMS. This enabled us to recapitulate and refine prior genotyping and methylation results, as well as to make several novel findings. By generating accurate consensus sequences for full-length patient alleles, we were able to perform detailed analysis of D4Z4 genetic variation, including the identification of haplotype-specific SNVs, and characterisation of inter-repeat and inter-array variation. Moreover, we discovered a previously-unreported 4qA variant, ‘4qAM’, which differs from the previously-identified 4qAS and 4qAL variants by the length of its distal repeat unit, but has an identical pLAM sequence and intact polyadenylation site. Further studies could elucidate the prevalence and distribution of 4qAM alleles in the general population, whether they can serve as the causative allele for FSHD, and whether they give rise to different *DUX4* transcripts, as is the case for 4qAS and 4qAL alleles (22). If 4qAM alleles are indeed permissive for *DUX4* expression, this would need to be considered in FSHD genetic testing, including updated conversion tables for calculation of repeat unit size using southern blotting, and potentially 4qAM-specific primers in addition to the 4qAS- and 4qAL-specific primers already used for (bisulfite)-PCR assays (15, 22, 38).

Accurate haplotyping and the capture of full-length spanning reads also allowed us to analyse the allele-specific and array-wide methylation profiles of FSHD and BAMS D4Z4 alleles, in aggregate and at single-molecule resolution. We were able to observe broad patterns in D4Z4 methylation that correlate with D4Z4 array length and *SMCHD1* status, including clear hypomethylation of contracted arrays in FSHD1, and of all 4q and 10q arrays in FSHD2. Notably, one of the samples that had been clinically-diagnosed as FSHD2 (34140) displayed comparatively high levels of methylation for all alleles, consistent with the absence of contracted arrays or pathogenic *SMCHD1, DNMT3B* or *LRIF1* variants. This illustrates that our workflow can be used to confirm a diagnosis of FSHD in patients with clinical manifestations of the disease, or conversely, to suggest an alternative diagnosis.

High-coverage and full-length methylation profiles also yielded several interesting findings. Many arrays showed a stepwise proximal-to-distal increase in methylation, however we observed some arrays with ‘atypical’ methylation patterns, especially in intermediate-length fibroblast alleles. Of note, overall DNA methylation patterns were highly consistent between individual molecules from the same allele, even for arrays with atypical methylation patterns, although not at the level of individual CpG sites. This aligns with observations from previous studies, which have shown that overall DNA methylation patterns can be stably inherited in clonal populations while being heterogeneous at the per-base level, which may represent differences in nucleosome phasing between molecules, stochasticity in the *de novo* and maintenance activities of epigenetic modifiers, and the influence of the methylation states of neighbouring CpGs (62, 65, 66). Future work should reveal whether those full-length D4Z4 methylation characteristics hold in other relevant cell types, such as myocytes to investigate disease pathogenesis and primary peripheral blood cells which could be used for diagnostic testing.

Previous studies have also indicated that BAMS is associated with D4Z4 hypomethylation in a similar manner to in FSHD2 (31), and here we have performed the first full-length analysis of methylation at the D4Z4 array in BAMS patients. Our results confirm that, at least in primary fibroblasts, D4Z4 methylation patterns are highly similar between BAMS and FSHD2, with retention of oscillatory patterns at the ~3.3 kbp and ~180 bp level, but with overall marked hypomethylation. So far, there has only been one reported case of a patient with overlapping symptoms of BAMS and FSHD, on the background of a FSHD2-permissive D4Z4 genotype (31). Shaw et al. (31) and Mohassel et al. (67) identified several other BAMS patients without clinical FSHD who had potentially permissive D4Z4 genotypes, however array size information was not available for all patients, D4Z4 methylation values were aggregate results obtained via bisulfite sequencing, and for young patients FSHD onset might not have been reached (as it typically manifests in young adults). Further genotyping and allele-specific methylation analysis could be used to more comprehensively characterise FSHD-permissive alleles in BAMS patients, and explore the possibility of as-yet undiscovered mechanisms that protect these patients from FSHD.

Finally, we also characterised several complex D4Z4 alleles containing duplicated D4Z4 arrays. Our results suggest that 4qA, 4qB and 10qA duplication alleles likely arise via the same mechanism, which results in in-*cis* duplications of the distal end of the subtelomere in the non-inverted orientation. We found that downstream in-*cis* duplicated arrays are highly methylated, whereas upstream inverted D4Z4 repeats are hypomethylated, which may suggest orientation-dependent long-range DNA methylation spreading mechanisms, inheritance of methylation patterns from the ancestral array, or insulator mechanisms that generate local differences in chromatin organisation. Lemmers et al. (45) found that in-*cis* duplication alleles may be able to give rise to FSHD-level *DUX4* expression and cause clinical FSHD in the absence of D4Z4 hypomethylation, demonstrating the need to further characterise the epigenetic mechanisms mediating *DUX4* repression.

We anticipate that, as a combined assay for D4Z4 haplotyping, D4Z4 methylation, and FSHD2-associated gene genotyping, our D4Z4End2End workflow has the potential to greatly simplify and streamline FSHD diagnostics. Several adaptations to the protocol could optimise it even further for use in clinical testing. First, in our study we use a single flow cell per patient sample, however we envisage that real-time analysis could facilitate dynamic processing of the sequencing data using the haplotyping pipeline, so that runs can be stopped once enough spanning reads are reached. This could allow multiple samples to be run on a single flow cell after sequential washes and reloads, decreasing sequencing costs. Second, adaptive sampling has recently emerged as an alternative to Cas9 for targeted sequencing (68), and may be able to further simplify library preparation while also capturing sequences further upstream and downstream of the D4Z4 array, obviating the need for separate whole-genome sequencing. As adaptive sampling targets regions of interest based on an easily-editable browser extensible data (BED) file, another benefit of this approach is that, with no added cost or complexity in library preparation, it could allow additional genetic loci to be added with the discovery of new causative variants for FSHD, as well as enabling optional inclusion of loci responsible for other neuromuscular diseases (NMDs). Indeed, previous studies have demonstrated the efficacy of adaptive sampling for diagnosing short-tandem repeat (STR) expansion-associated NMDs (69, 70), and we imagine that STRs, the D4Z4 region, FSHD2-associated genes, and causative loci for other genetic NMDs could be combined into a single neuromuscular gene panel for targeted sequencing.

Moreover, there are still many unknowns surrounding D4Z4 regulation, and our approach could serve as a powerful research tool to answer fundamental questions in macrosatellite biology, such as further investigating the dynamic changes in D4Z4 methylation across different cell types, stages of development, and disease states. Indeed, in addition to embryonic development and FSHD, *DUX4* expression is also seen physiologically in the human testis and thymus, and pathologically in several solid cancers, inflammatory disorders, and herpesvirus infection (7). Elucidating the epigenetic changes in these contexts may help to reveal the underlying mechanisms of *DUX4* silencing and reactivation, paving the way for the identification of new therapeutic targets. Additionally, with the recent development of simultaneous profiling techniques such as DiMeLo-seq (71), nanoNOMe (72) and nanoHiMe-seq (73), the long-read, single-molecule approach presented here has the potential to be extended to the exploration of even more dimensions of D4Z4 regulation, such as SMCHD1 and CTCF binding, nucleosome occupancy, and histone modifications, alongside genetic sequence and methylation. Our workflow for investigating macrosatellite (epi)genetic landscapes therefore provides a means to explore alterations in the molecular machinery in functional studies, investigate the effects of emerging gene therapies, and map these dynamic and intriguing repetitive regions of the genome with much more depth, detail and precision than previously possible.

## Methods

### Patient sample collection and cell culture

All participants provided signed informed consent for research and publication at the time of recruitment. The study complied with the Declaration of Helsinki. This study received ethics approval from the Walter Eliza Hall Institute (Ethics ID: 20/16B) and the French Agency of Biomedicine (CRBAPHM OCP05P01E003; Étude D-2023-FSHD). FSHD and BAMS patients were clinically diagnosed and have previously received clinical genetic testing for D4Z4 array size (FSHD and controls), D4Z4 methylation (FSHD) and *SMCHD1* genotype (FSHD, BAMS and controls), as described in Supplementary Table S3. Controls were previously confirmed to have D4Z4 array size >10 units and to have no pathogenic *SMCHD1* variant. Primary fibroblasts from FSHD and BAMS patients and controls were obtained via skin biopsies. Fibroblasts were cultured in DMEM with 4.5 g/L D-glucose, FBS South American, PenicillinStreptomycin 5000 U/mL, and GlutaMAX™ (Thermofisher), and incubated at 37°C, 5% CO2 / 9% air in a humidified incubator.

### DNA extraction

DNA was extracted from primary fibroblasts using the Monarch® high-molecular weight (HMW) DNA extraction kit for cells and blood as per the protocol (NEB T3050) using 300 rpm agitation, and eluted in elution buffer (EB) from the relevant Oxford Nanopore Technologies (ONT) kit used for library preparation (SQK-ULK001, SQK-ULK114 or SQK-CS9109).

### Nanopore library preparation and sequencing

For ultra-long whole-genome sequencing, libraries were prepared using the ONT ultra-long DNA sequencing kit (either SQK-ULK001 or SQK-ULK114) according to the relevant ONT protocol. For Cas9-targeted sequencing, Cas9 crRNAs targeting 4q/10q, *SMCHD1, DNMT3B* and *LRIF1* (Supplementary Table S3) were designed using *CHOPCHOP* (v3) (GRCh38 or CHM13 T2T v1.1, CRISPR/Cas9, nanopore enrichment, ‘Doench et al. 2014 – only for NGG PAM’).

Cas9 RNP formation and DNA library preparation for Cas9-targeted sequencing were performed using the ONT Cas9 Sequencing Kit (SQK-CS9109) as per the ONT protocol. Libraries were loaded on R9.4.1 PromethION flow cells (FLO-PRO002) if prepared using SQK-ULK001 or SQK-CS9109, and on R10.4.1 (FLO-PRO114M) if prepared using SQK-ULK114, and sequenced to exhaustion on a PromethION P24.

### Basecalling

Reads from fast5 files were basecalled using the latest *Guppy* version at the time of sequencing (v6.1.1 or v6.3.4) using the modified basecalling model dna_r9.4.1_450bps_modbases_5mc_cg_sup_prom.cfg.

Reads from pod5 files from later sequencing runs were basecalled using the latest *Dorado* version at the time of sequencing (v0.3.2 or v0.3.3), using model dna_r10.4.1_e8.2_400bps_sup@v4.2.0 with the argument `--modified-bases 5mCG_5hmCG` for concurrent modified basecalling.

### Haplotyping pipeline

Processing of fastq files to identify, annotate and haplotype 4q and 10q reads was performed using a custom script, available at https://github.com/lucindaxiao/D4Z4End2End. Briefly, reads were mapped against a D4Z4 reference sequence (the first full KpnI-KpnI D4Z4 unit from the 4q array of CHM13v1.0) using *minimap2* (v2.24), to identify D4Z4-containing reads and count the number of D4Z4 units. These reads were then mapped against the 4q and 10q regions of CHM13v2.0, and secondary and supplementary alignments were filtered out using `samtools view -F 2304`, to identify 4q- and 10q-specific D4Z4 reads and assign them to 4q or 10q haplotypes. D4Z4-containing reads were also mapped against reference sequences for p13E-11 (from (74)), pLAM (sequence from end of last D4Z4 to end of *DUX4* exon 3 in 4q allele of CHM13v2.0), and the 4qB distal region (from GRCh38) for read annotation. Reads containing pLAM and qB-sequence were assigned to haplotype A and B, respectively. Reads containing both p13E-11 and a D4Z4-distal feature (pLAM or qB-sequence) were classified as spanning reads, and all other reads were classified as non-spanning reads. Reads were also searched for exact matches for XapI sites, BlnI sites, and 4qA/10qA-specific poly(A)-signal sequences using regular expression matching in Python. *Noise-Cancelling Repeat Finder* (v1.01.00) (75) was used to search reads for beta-satellite repeats (from (76)) and telomeric repeats. The interactive visualisation tool for exploration of haplotyped reads was created using *D3*.*js*. The haplotyping pipeline was used to process the ultra-long whole-genome sequencing and Cas9-targeted sequencing data from all patient samples, and publicly-available ONT sequencing data for CHM13v2.0 from the Telomere to Telomere (T2T) Consortium (accessed from s3://human-pangenomics/T2T/CHM13/nanopore/rel8-guppy-5.0.7/reads.fastq.gz) and for five cell lines from the Human Reference Pangenome Consortium (HPRC) (accessed from s3://human-pangenomics/working/HPRC/).

### D4Z4 genetic analysis

For comparison of the haplotyping pipeline results for CHM13 and the HPRC cell lines to their corresponding assemblies (CHM13v2.0 and HPRC assemblies), D4Z4 arrays in the assemblies were annotated by mapping against the D4Z4, p13E-11, pLAM and 4qB reference sequences described above, using *minimap2* (77).

Consensus sequences for patient 4q and 10q alleles were generated from spanning reads identified by the haplotyping pipeline, using a two-step process: (1) *minimap2* (77) for overlap detection, followed by (2) *Racon* (v1.4.20) (25) for consensus calling, using the `--no-trimming` argument and using the longest spanning read as the target sequence for correction.

*Minimap2* (77) was used to extract 4q/10q D4Z4 units from the consensus sequences and from the CHM13v2.0 and GRCh38 assemblies (GRCh38 accessed from http://hgdownload.soe.ucsc.edu/goldenPath/hg38/chromosomes/), and to map them against a 4qA-type D4Z4 reference sequence. The 4qA-type D4Z4 reference sequence was generated by performing a *MUSCLE* (v3.8) (78) multiple sequence alignment of all full-length D4Z4 units from the CHM13v2.0 4q allele, and taking the most frequent value at each position (nucleotide or gap). D4Z4 alignments were visualised using *Integrative Genomics Viewer* (v2.17.4) (79).

*MUSCLE* was also used for multiple sequence alignments of distal D4Z4 sequences, which were visualised using the *R* package *ggmsa* (80).

For the BAMS1 sample, *WhatsHap* (v2.1) (81) was used to phase the non-spanning 4qA reads that overlapped the proximal end of the D4Z4 array.

### FSHD2 gene panel variant calling

BAM files for patient samples were run through *DeepVariant* (v1.5.0) (23) using model type ONT_R104, to generate variant call format (VCF) files targeting *SMCHD1, DNMT3B* and *LRIF1*. VCF files were filtered for ‘PASS’ calls using bcftools (82). VCF files were input into *Variant Effect Predictor (VEP)* (v107.0) (24) using the T2T-CHM13v2.0 *VEP* cache file, to annotate variant calls for potential functional impact. Variant calls overlapping *SMCHD1, DNMT3B* or *LRIF1* with IMPACT=HIGH or IMPACT=MODERATE as output by *VEP* were further assessed by comparison to previously reported pathogenic *SMCHD1* variants (https://databases.lovd.nl/shared/genes/SMCHD1) and variants in healthy controls, as well as variant pathogenicity predictions from *AlphaMissense* (59).

### Methylation analysis

To generate allele-specific methylation plots, haplotyped reads were extracted from the mod-BAM files and re-aligned using *minimap2* to either the allele’s *Racon* consensus sequence, or a raw read from the allele if no consensus sequence was generated. For samples with high coverage Cas9-targeted sequencing (17706, 19187, BAMS9), all 4q and 10q reads were mapped against the full set of consensus sequences for the sample’s alleles, to haplotype both spanning and non-spanning reads. For samples with lower coverage, reads included for methylation analysis were either spanning reads, or reads that could be confidently assigned to an allele based on allele-specific differences in SNVs (XapI or BlnI sites for 4q vs 10q), distal sequences (AS/AM/AL/B haplotype) and/or number of repeat units. Allele-specific methylation plots were generated using *NanoMethViz* (26, 27), using the `plot_region()` function with `smoothing_window=2000`. Annotations shown below the methylation plots were generated using *minimap2*, as described above.

To perform allele-specific autocorrelation of 5mC signals, bedmethyl files were first produced from the re-aligned modBAM files using *modkit* (v0.2.5) (https://github.com/nanoporetech/modkit) with the `--cpg` and `--combine-strands` arguments. A vector of length *n* where *n* is the length in nucleotides of the reference sequence for the allele was populated with the %5mC (‘fraction modified’ from *modkit*) for all CpG sites with coverage ≥5, and ‘NA’ otherwise. The `acf()` function within *R* was then used to calculate the autocorrelation between %5mC values across the length of the vector, and the results were plotted for lag values (corresponding to distance in nucleotides between CpG sites) of up to 10000 nucleotides (approximately three full D4Z4 units) and up to 500 nucleotides. Allele-specific %5mC plots were generated from the bedmethyl files, once again filtering for CpG sites with coverage ≥ 5, using *ggplot2* (v3.5.1) (83). Smoothed lines were generated using the `geom_smooth()` function using the LOESS method with `span = 0.2`.

Mean methylation rate was calculated using the `region_methy_stats()` function within *NanoMethViz* (26, 27), using a threshold of 0.5 to call methylated and unmethylated sites. Regression lines for the scatterplots of mean methylation prevalence vs number of repeats were produced using `geom_line(stat=“smooth”, method = “lm”)`, and Pearson and Spearman correlations were calculated using the `cor()` function within *R*. For comparison of the mean methylation prevalence of the DR1 site and *DUX4* transcription start sites (TSS) within each repeat unit, the DR1 site was defined as nucleotides 563–814 of the D4Z4 unit with respect to the start of the KpnI site (37), and the *DUX4* TSS region was defined as −200nt to +200nt with respect to the *DUX4* TSS (nucleotide 10732 in Genbank AF117653.3).

## Data Availability

1KGP-ONT data are available on https://s3.amazonaws.com/1000g-ont/index.html. Human Pangenome Consortium draft assemblies and data used are available on https://github.com/human-pangenomics/HPP_Year1_Assemblies.

https://github.com/human-pangenomics/HPP\_Year1\_Assemblies

## Data access

1KGP-ONT (63) data available on https://s3.amazonaws.com/1000g-ont/index.html. Human Pangenome Consortium draft assemblies and data used are available on https://github.com/humanpangenomics/HPP_Year1_Assemblies.

## Acknowledgements

We thank Stephen Wilcox and Sarah MacRaild from the WEHI Genomics Platform for their assistance with nanopore sequencing; Kelsey Breslin and Hannah Vanyai for their help with cell culture.

