## Supplemental tables and figures for "D4Z4End2End: complete genetic and epigenetic architecture of D4Z4 macrosatellites in FSHD, BAMS and reference cohorts"

| | Patient samples<br>(# of individuals) | Targeted/<br>WGS approach | Alleles studied | | | Methylation | Mosaicism,<br>complex alleles<br>(# of alleles) | FSHD2<br>genes |
|---|---|---|---|---|---|---|---|---|
| | | | Hap. | Range of array size (# of alleles) | Max. # of spanning reads for a single allele | | | |
| *Hiramuki et al., 2022* [1] | FSHD1 (12)<br>FSHD2 (2) | Cas9-targeted<br>- p13E-11, A-sequence | 4qA<br>10qA | 1-13 RU (15)<br>7-13 RU (9) | 15 (2RU)<br>6 (8RU) | Per-RU for 4qA and 10qA reads; promoter + gene body for distal RU | N/A | N/A |
| *Yeetong et al., 2023* [2] | FSHD1 (7)<br>Unaffected parents (6)<br>Controls (10) | WGS | 4qA<br>10qA | 1-5 RU (7)<br>6-9 RU (3) | 6 (2RU)<br>3 (6RU) | N/A | N/A | N/A |
| *Butterfield et al., 2023* [3] | FSHD1 (8)<br>FSHD2 (2)<br>*Controls (1)** | Cas9-targeted<br>- p13E-11, pLAM<br>- D4Z4<br><br>*Publicly-available WGS** | 4qA<br><br>10qA<br><br>*4qB** | 2-25 RU (11)<br>*42 RU (1)**<br>7-24 RU (12)<br>*20-21 RU (2)**<br>*16 RU (1)** | 78 (5 RU)<br><br>63 (7 RU) | Base-level and per-RU for 4qA, 10qA and 4qB alleles | N/A | N/A |
| *Li et al., 2024* [4] | FSHD1 (1) | WGS | 4qA | 5 RU (1) | 3 (5 RU) | N/A | N/A | N/A |
| *Lemmers et al., 2024* [5] | FSHD1 (2)<br>Unaffected mother (1) | Cas9-targeted<br>- p13E-11, pLAM | 4qA | 17+2 RU (1)<br>17+9 RU (1)<br>2+10 RU (1) | Spanning reads obtained for individual arrays and spacer, but none spanning both arrays | Base-level and per-RU for 4qA *in-cis* duplication alleles | Duplication (3) | N/A |
| *Huang et al., 2024* [6] | FSHD1 (12)<br>Controls (11)<br>iPSCs (6) from FSHD1 and controls | WGS | 4qA<br>4qB<br>10qA | 2-21 RU (23)<br>7-20 RU (7)<br>6-23 (21) | 8 (4RU)<br>4 (7RU)<br>10 (6RU) | Base-level and average for distal RU for 4qA alleles | Mosaicism (1) | *SMCHD1*,<br>*DNMT3B*,<br>*LRIF1* |
| *Wang et al., 2024* [7] | FSHD1 (1) – prenatal diagnosis via amniocentesis | WGS | 4qA<br>4qB | 4 (1)<br>8 (1) | 2 (4RU)<br>2 (8RU) | Per-RU for 4qA and 4qB alleles | N/A | N/A |
| ***Our study*** | FSHD1 (4)<br>FSHD2 (4)<br>BAMS (2)<br>Controls (2) | Cas9-targeted<br>- 4q and 10q<br>- FSHD2 gene panel<br><br>WGS | 4qA<br>4qB<br>10qA<br>10qB | 1-42 RU (14)<br>13-26 RU (5)<br>6-36 (20)<br>13-19 (3) | 116 (2RU)<br>79 (13RU)<br>163 (7RU)<br>3 (17RU) | Base-level and per-RU for 4qA, 4qB, 10qA, 10qB alleles (incl. duplicated arrays and upstream inverted RUs) | Mosaicism (2)<br>Duplication (2)<br>Triplication (1)<br>Upstream inverted array (1) | *SMCHD1*,<br>*DNMT3B*,<br>*LRIF1* |

**Supplementary Table S1. Comparison of our study with previous Nanopore-based studies for FSHD.** Our study extends upon previous studies by analysing samples from BAMS patients, including guide RNAs for FSHD2-associated genes for Cas9-targeted sequencing, capturing longer full-length D4Z4 alleles with higher coverage, and analysing the structure and methylation of full-length complex alleles, including those with duplications and triplications of the D4Z4 array.

| | Haplotypes from Nanopore raw spanning reads (full D4Z4 units) | HPRC assembly | | | | |
|---|---|---|---|---|---|---|
| | | Scaffold | p13E11 | pLAM/ B-sequence | No. of full D4Z4 units Start / middle / end / full array | |
| **HG00621** | 4qAS 13 RU<br>4qB 15 RU<br>10qA 10 RU<br>10qA 19 RU | HG00621#2#JAHBCC010000040.1 | Y | -- | 8 | start |
| | | HG00621#2#JAHBCC010000132.1 | Y | pLAM | 19 | full |
| | | HG00621#2#JAHBCC010000194.1 | -- | B-sequence | 12 | end |
| | | HG00621#1#JAHBCD010000073.1 | Y | pLAM | 13 | full |
| | | HG00621#1#JAHBCD010000149.1 | Y | pLAM | 10 | full |
| **HG00735** | 4qAS 34 RU<br>4qB 46 RU<br>10qA 4 RU<br>10qA 31 RU | HG00735#2#JAHBCG010000093.1 | Y | -- | 11 | start |
| | | HG00735#2#JAHBCG010000109.1 | Y | pLAM | 4 | full |
| | | HG00735#2#JAHBCG010000195.1 | -- | B-sequence | 18 | end |
| | | HG00735#2#JAHBCG010000230.1 | -- | -- | 15 | middle |
| | | HG00735#1#JAHBCH010000030.1 | Y | -- | 6 | start |
| | | HG00735#1#JAHBCH010000093.1 | Y | -- | 14 | start |
| | | HG00735#1#JAHBCH010000168.1 | -- | pLAM | 5 | end |
| | | HG00735#1#JAHBCH010000258.1 | -- | pLAM | 7 | end |
| **HG00741** | 4qB 10 RU<br>4qAS 38 RU<br>10qA 20 RU | HG00741#2#JAHALX010000140.1 | Y | B-sequence | 10 | full |
| | | HG00741#2#JAHALX010000136.1 | Y | -- | 11 | start |
| | | HG00741#2#JAHALX010000243.1 | -- | pLAM | 6 | end |
| | | HG00741#1#JAHALY010000013.1 | Y | -- | 10 | start |
| | | HG00741#1#JAHALY010000157.1 | -- | pLAM | 5 | end |
| | | HG00741#1#JAHALY010000230.1 | Y | -- | 4 | start |
| | | HG00741#1#JAHALY010000244.1 | -- | pLAM | 8 | end |
| **HG01106** | 4qB 15 RU<br>4qA 28 RU<br>10qA 17 RU | HG01106#2#JAHAMB010000127.1 | Y | -- | 6 | start |
| | | HG01106#2#JAHAMB010000188.1 | -- | B-sequence | 9 | end |
| | | HG01106#2#JAHAMB010000214.1 | -- | pLAM | 5 | end |
| | | HG01106#1#JAHAMC010000060.1 | Y | -- | 7 | start |
| | | HG01106#1#JAHAMC010000067.1 | Y | -- | 7 | start |
| | | HG01106#1#JAHAMC010000178.1 | -- | pLAM | 18 | end |
| | | HG01106#1#JAHAMC010000192.1 | -- | pLAM | 5 | end |
| **HG01175** | 4qB 33 RU<br>4qAS 52 RU<br>10qA 8 RU<br>10qB 20 RU | HG01175#2#JAHALZ010000099.1 | Y | pLAM | 3 | full |
| | | HG01175#2#JAHALZ010000122.1 | Y | -- | 3 | start |
| | | HG01175#2#JAHALZ010000265.1 | -- | pLAM | 8 | end |
| | | HG01175#1#JAHAMA010000091.1 | Y | -- | 6 | start |
| | | HG01175#1#JAHAMA010000128.1 | Y | B-sequence | 9 | full |
| | | HG01175#1#JAHAMA010000263.1 | -- | B-sequence | 20 | end |

**Supplementary Table S2. Comparison of D4Z4 genotypes determined using raw Nanopore sequencing data to draft assemblies from the Human Pangenome Reference Consortium (HPRC).** The haplotyping pipeline was used to process publicly-available raw Nanopore sequencing data generated by the HPRC[8] for 5 lymphoblastoid cell lines (LCLs) from the 1000 Genomes Project[9]. 4q and 10q haplotypes were able to be determined from spanning reads present in the raw data. D4Z4 arrays within the corresponding HPRC draft assemblies were then assessed for their number of repeat units, and for whether they spanned the full D4Z4 array.

| SAMPLE | Sex | D4Z4 haplotypes | *SMCHD1* genotype | %5mC DR1 |
|---|---|---|---|---|
| **Control** | | | | |
| **AG09309** | F | >10 | No mutation | Not known |
| **AG10803** | M | >10 | No mutation | Not known |
| **FSHD1** | | | | |
| **12566**[10,11] | F | 4qA 3RU | No mutation | 36.8% |
| **FSHD1_3 (TalF[10,11], 37-$I_1$[12])** | M | 4qA 2RU<br>4qB 18RU<br>10qA 36RU<br>10qA 15+2+6RU (cis-triplication)[12] | No mutation | 45.8% |
| **17706**[10,11] | F | 4qA 2RU (25%)<br>4qA 15RU<br>4qA 37RU<br>10qA 6RU<br>10qA 14RU | No mutation | 38.6% |
| **19187** | F | 4qA 2RU (50%)<br>4qA 22RU<br>4qA 32RU<br>10qA 14RU<br>10qA-cis duplication | No mutation | 45.7% |
| **FSHD2** | | | | |
| **11440**[13] | M | 4qA 11RU<br>4qA 34RU<br>10qA 15RU<br>10qA 17RU | c.2338+4A>G | 16.4% |
| **15166**[13] | F | 4qA 11RU<br>4qB 21RU<br>10qB 13RU<br>10qB 20RU | c.4608_4614dup<br>p.Ala1539Tyrfs*6 | 11.3% |
| **34140** | F | 4qA 29RU<br>4qA 30RU<br>10qA 17RU<br>10qA 19RU | p.L1031 | 37% |
| **11491**[13] | F | 4qA 22RU<br>4qA 22RU<br>10qA 6RU<br>10qA 6RU | c.5476+3A>G | 9.8% |
| **BAMS** | | | | |
| **BAMS1**[14] | M | Not known | c.407A>G<br>p.E136G | Not known |
| **BAMS9**[14] | M | Not known | c.1259A>T<br>p.D420V | Not known |

**Supplementary Table S3. Previous genotyping and methylation data for control, FSHD and BAMS fibroblasts.** FSHD and BAMS patients were diagnosed based on clinical findings and clinical genetic testing. D4Z4 genotyping was performed via Southern Blot and/or molecular combing. Methylation analysis was performed via bisulfite sequencing using PCR primers specific for the DR1 region.

| | |
|---|---|
| D4Z4_up_1 | GATACCGACAGCAATAGTCC |
| D4Z4_up_2 | GTTGTGAAGTTAGAAGGTGC |
| D4Z4_A_down_1 | GTATGCTGCGGGTTGTGGGG |
| D4Z4_A_down_2 | GAACACACTACCTTTCCATG |
| D4Z4_B_down_1 | GGAATGTATAATACTTCTGC |
| D4Z4_B_down_2 | GAGTCTACAGTAGTGTTCGA |
| smchd1_p1_up | CCAACTTCGCGAGGGCCGAG |
| smchd1_p1_down | AGGAGATCGAACATGACAAC |
| smchd1_p2_up | CTAAGCACCCTACTTTAATT |
| smchd1_p2_down | GAGCAGTGCAAGAGTGAAAG |
| smchd1_p3_up | CATTGACCCATACTTCGAGA |
| smchd1_p3_down | TTAGGAGATATCATTTACGA |
| smchd1_p4_up_1 | TTACCTTCTTAAGCAGTGCA |
| smchd1_p4_up_2 | GTGATTTCATCCTTTGACCA |
| smchd1_p4_up_3 | ACACTATTTGTACTCTCTTG |
| smchd1_p4_down | AATGATAACCCACTGCCATA |
| dnmt3b_exon1_up | CCTGTCCTTAGTTTACTGCG |
| dnmt3b_exon1_down | GCAATGAACTTGGCGAGACA |
| dnmt3b_rest_up | GTGAAAATCCCGCTTCAGGC |
| dnmt3b_rest_down | GGGCTTGCCCGTCTGTCTTA |
| lrif1_up | ACTCACTTAAAAGCTCTACA |
| lrif1_down | GACTAATTAATAGTGCTGCG |

**Supplementary Table S4. Guide RNAs (gRNAs) used for Cas9-targeted sequencing.** gRNAs were designed using CHOPCHOP v3[15] (GRCh38 or CHM13 T2T v1.1, CRISPR/Cas9, nanopore enrichment, 'Doench et al. 2014 – only for NGG PAM').

| Cell line | 4q | | 10q | |
|---|---|---|---|---|
| | Hap. | RUs | Hap. | RUs |
| **HG00099** | 4qB | 23 | 10qA | 7 |
| | 4qB | 2.5* + 35 | -- | -- |
| **HG00105** | 4qAS | 37 | 10qA | 22 |
| | -- | -- | 10qA | 27 |
| **HG00110** | 4qB | 23 | 10qA | 16 |
| | 4qAL | 30 | -- | -- |
| **HG00121** | 4qAL | 18 | 10qA | 14 |
| | -- | -- | -- | -- |
| **HG00127** | 4qB | 19 | 10qA | 11 |
| | -- | -- | 10qA | 31 |
| **HG00136** | 4qB | 17 | 10qA | 15 |
| | -- | -- | 10qA | 30 |
| **HG00151** | 4qAS | 31 | 10qA | 12 |
| | -- | -- | 10qA | 31 |
| **HG00410** | 4qB | 12 | 10qA | 21 |
| | 4qAS | 29 | -- | -- |
| **HG00525** | -- | -- | 10qA | 34 |
| | -- | -- | 10qA | 41 |
| **HG00631** | 4qAS | 13 | 10qA | 7 |
| | -- | -- | 10qA | 19 |
| **HG00675** | 4qAS | 21 | 10qA | 10 |
| | 4qB | ≥18 + 3^ | 10qA | 13 |
| **HG00706** | 4qB | 24 | 10qA | 7 |
| | -- | -- | -- | -- |
| **HG00728** | 4qB | 13 | 10qA | 10 |
| | -- | -- | 10qA | 14 |
| **HG01046** | 4qB | 22 | 10qA | 25 |
| | 4qAS | 32 | -- | -- |
| **HG01122** | 4qB | 40 | 10qA | 23 |
| | -- | -- | 10qA | 9 |
| **HG01281** | 4qB | 30 | 10qA | 37 |
| | 4qAL | 26 | -- | -- |
| **HG01369** | 4qB | 23 | 10qA | 8 |
| | 4qB | 20 | -- | -- |
| **HG01372** | 4qAS | 21 | 10qA | 12 |
| | 4qAS | 31 | 10qA | 17 |
| **HG01395** | -- | -- | 10qA | 18 |
| | -- | -- | -- | -- |
| **HG01414** | 4qB | 26 | 10qA | 9 |
| | -- | -- | 10qA | 11 |
| **HG01468** | 4qAS | 27 | 10qA | 11 |
| | -- | -- | -- | -- |
| **HG01615** | -- | -- | 10qA | 22 |
| | -- | -- | 10qA | 15 |
| **HG01695** | 4qB | 20 | 10qA | 6 |
| | 4qAS | 26 | 10qB | 19 |
| **HG01790** | 4qB | 23 | 10qA | 38 |
| | 4qAS | 43 | -- | -- |
| **HG01797** | 4qB | 14 | 10qA | 7 |
| | 4qB | 28 | 10qA | 25 |
| **HG01801** | -- | -- | 10qA | 11 |
| | -- | -- | 10qA | 23 |
| **HG01812** | -- | -- | 10qA | 13 |
| | 4qAS | 26 | 10qA | 17 |
| **HG02185** | 4qAS | 26 | 10qA | 21 |
| | 4qB | 12 | 10qA | 24 |
| **HG02252** | 4qAS | 29 | 10qA | 10 |
| | -- | -- | -- | -- |
| **HG02282** | -- | -- | 10qA | 8 |
| | -- | -- | -- | -- |

*Upstream inverted D4Z4 array
^Downstream non-inverted D4Z4 array

**Supplementary Table S5. D4Z4 haplotypes for 30 B-lymphocyte lymphoblastoid cell lines (LCLs) from the 1000 Genomes Project[9].** Haplotypes were determined from spanning D4Z4 reads within publicly-available raw Nanopore sequencing data from the 1000 Genomes Project ONT Sequencing Consortium (1KGP-ONT)[16], as identified by processing the raw data using the haplotyping pipeline. Blank cells indicate that no spanning reads were identified for that allele.

***SMCHD1***

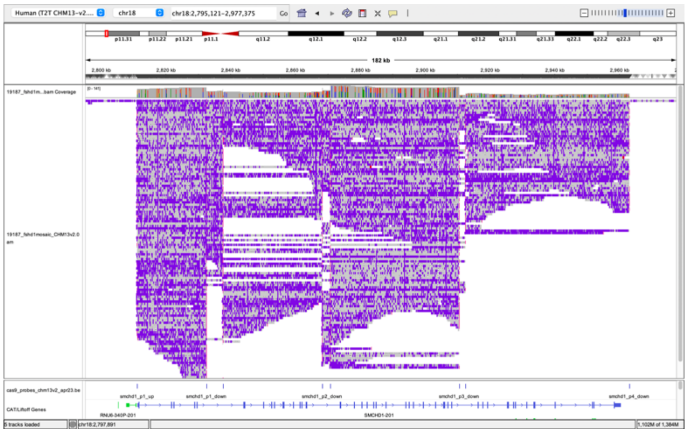

***DNMT3B***

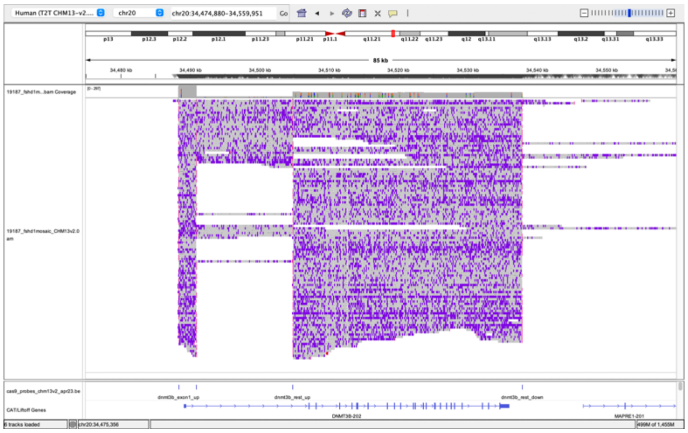

***LRIF1***

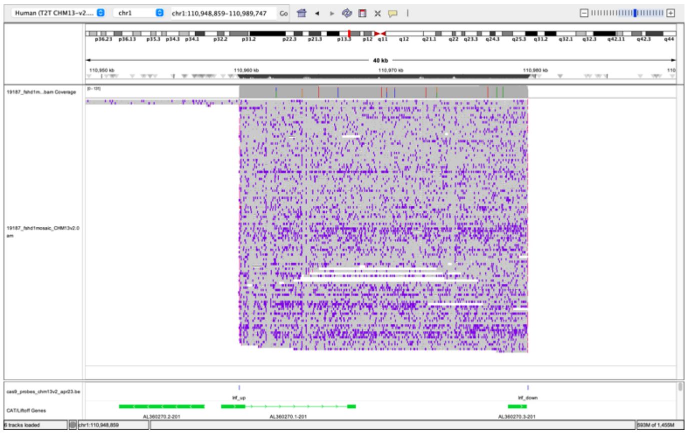

**Supplementary Figure S1. Coverage of FSHD2 gene panel using Cas9-targeted sequencing.** IGV screenshots of *SMCHD1*, *DNMT3B* and *LRIF1* coverage from Cas9-targeted sequencing of sample 19187 fibroblasts. Locations of Cas9 gRNAs (Supplementary Table S4) are shown below the alignments.

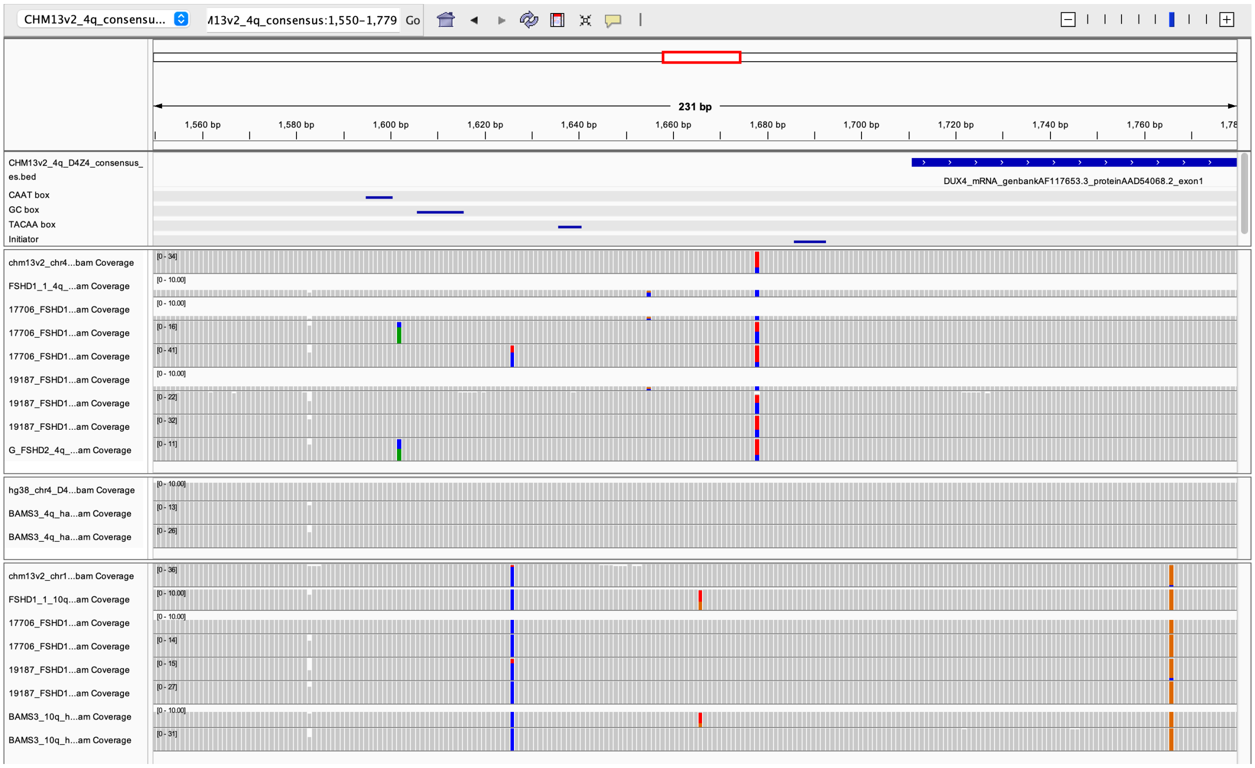


**Supplementary Figure S2. Sequence variants in the *DUX4* promoter region for 4qA, 4qB and 10qA alleles.** The locations of known *DUX4* promoter elements[17], are shown in the top panel, alongside the start of *DUX4* exon 1. Coverage tracks for alignments of D4Z4 units extracted from full-length D4Z4 consensus sequences are shown for 4qA alleles (second panel), 4qB alleles (third panel) and 10qA alleles (bottom panel), displaying 4qA-, 4qB- and 10qA-specific SNVs.

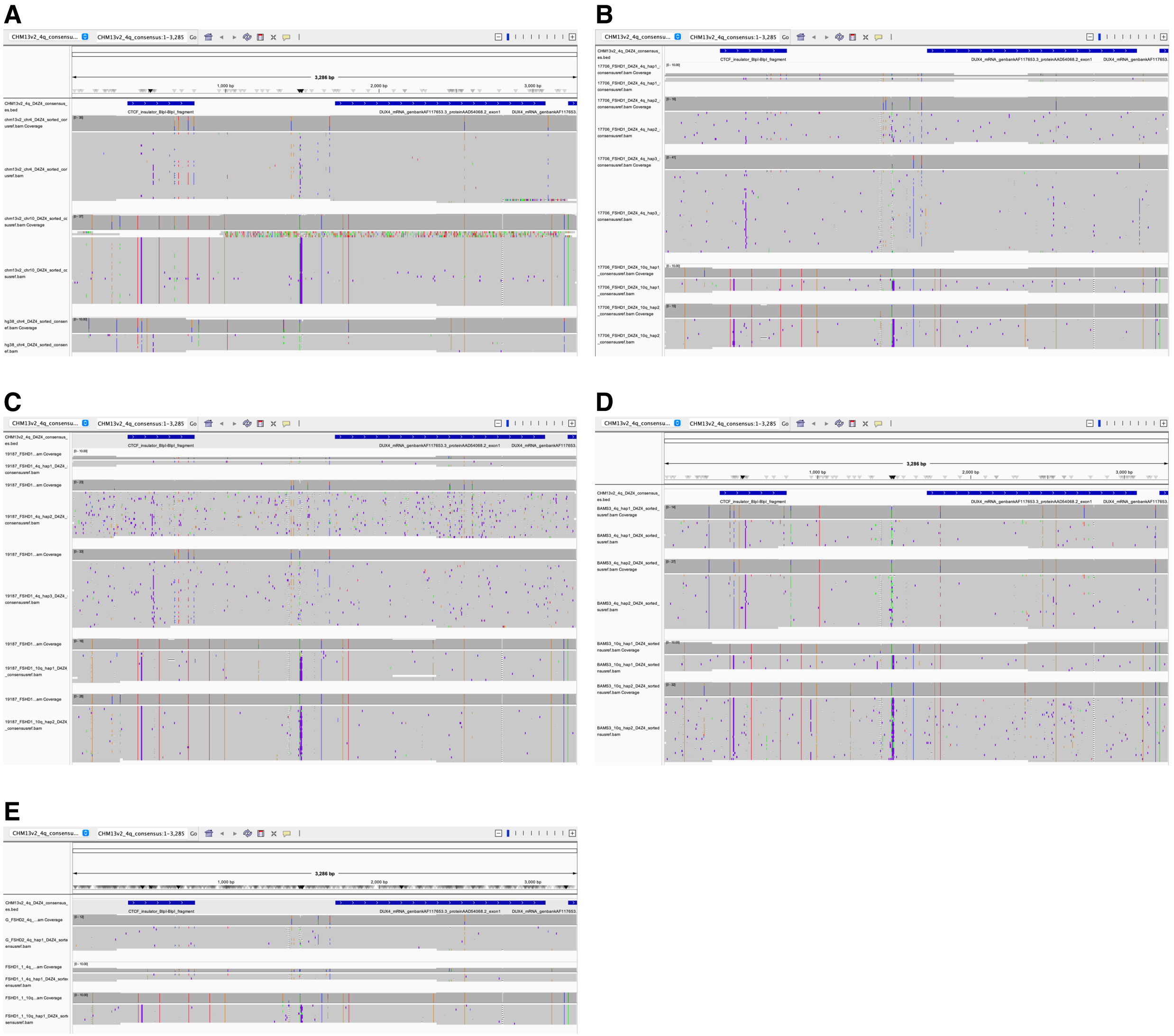

**Supplementary Figure S3. Analysis of inter-D4Z4 genetic variation for 4qA, 4qB and 10qA alleles.** Each panel shows alignments of individual D4Z4 units from the consensus sequences of full-length D4Z4 alleles, against a 4qA-type reference sequence (derived from CHM13v2.0, see Methods). Within each alignment track, full-length D4Z4 units are arranged from top to bottom in order of proximal to distal position within the array. D4Z4 units from (A) the 4qA and 10q arrays from CHM13v2.0, and the 4qB array from GRCh38 (B) sample 17706 (FSHD1, mosaic sample) (C) sample 19187 (FSHD1, mosaic sample); note that the 10qA 15RU allele ('10q_hap1') contains a truncated internal D4Z4 unit, and the proximal partial D4Z4 unit is shown in the same row as this truncated repeat (D) sample BAMS9 (E) samples 11440 (labelled as 'G_FSHD2') and 12566 (labelled as 'FSHD1_1').

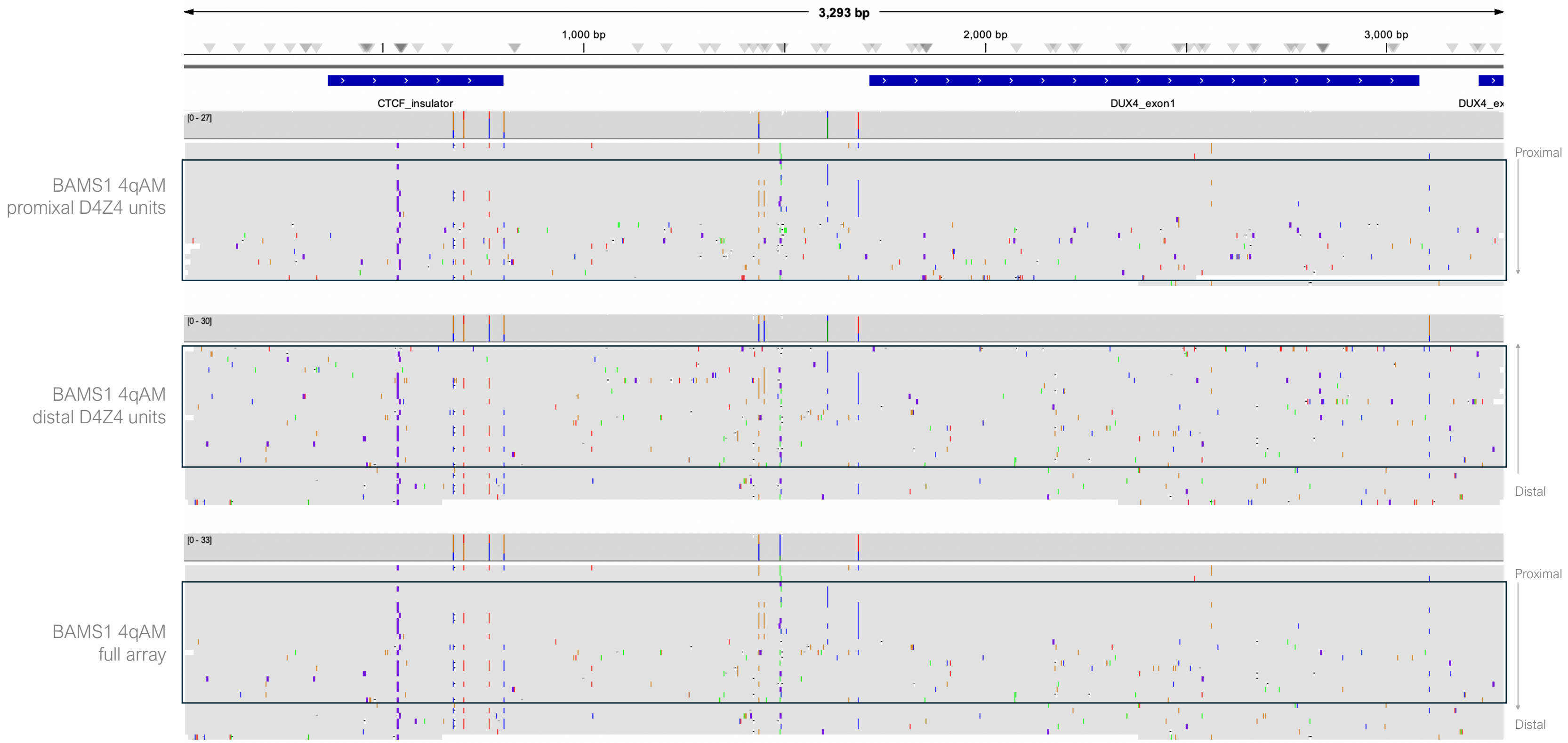


**Supplementary Figure S4. Allele-specific analysis of D4Z4 composition enables construction of full-length D4Z4 sequences from non-spanning reads.** No spanning 4q reads were obtained for the BAMS1 sample, however variants upstream of D4Z4 enabled 4q reads spanning the proximal end of the array to be phased into two haplotypes using WhatsHap[18], while reads spanning the distal end of the array could be phased based on their AS/AM genotype. Consensus sequences were then generated using Racon[19] for each of the groups of phased proximal (top panel) and distal (middle panel) reads. Based on the pattern of variants across the individual D4Z4 units, the region of overlap between the proximal and distal reads was able to be determined (black rectangles) to construct the sequence for the full-length D4Z4 array (bottom panel). The BAMS1 4qAM allele was found to have the same pattern of variants as the 19187_FSHD1 4qAM allele (Supplementary Figure S3C).

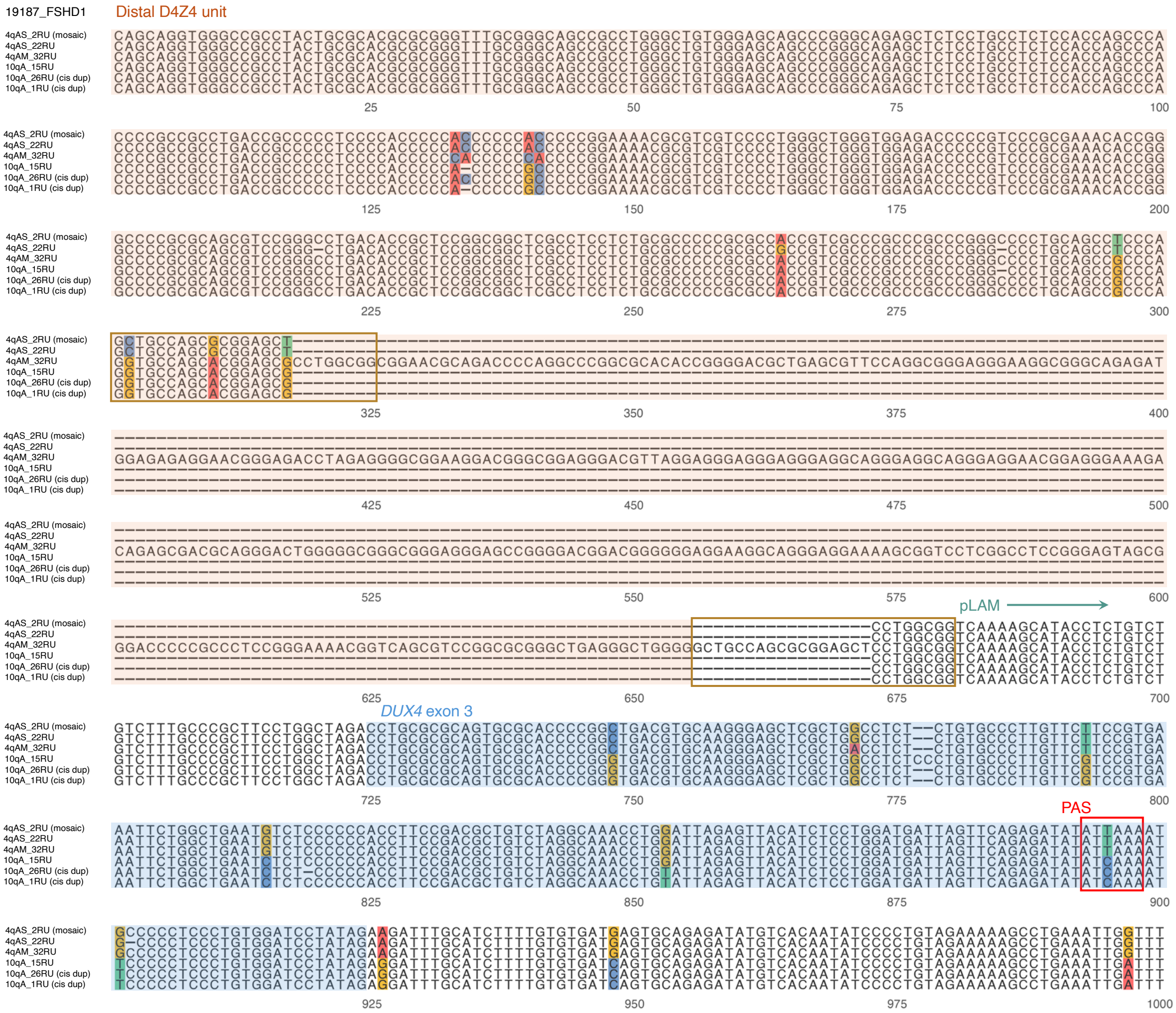


**Supplementary Figure S5. Alignment of distal D4Z4 sequences from 4q and 10q alleles from the mosaic FSHD1 sample 19187.** 19187 contains 4qAS, 4qAM and 10qA alleles, including an in-*cis* duplicated 10qA array which is also included in the alignment. The orange-boxed areas contain regions of homology between the 4qAM sequence immediately distal to the breakpoint for the final 4qAM D4Z4 unit, and a region within the D4Z4 sequence just proximal to the breakpoint for the 4qAS D4Z4 unit. Here the start of pLAM is marked as the first nucleotide following the final 4qAS D4Z4 unit. Sequences are extracted from consensus sequences generated using Racon[19] from high-coverage Cas9-targeted Nanopore sequencing.

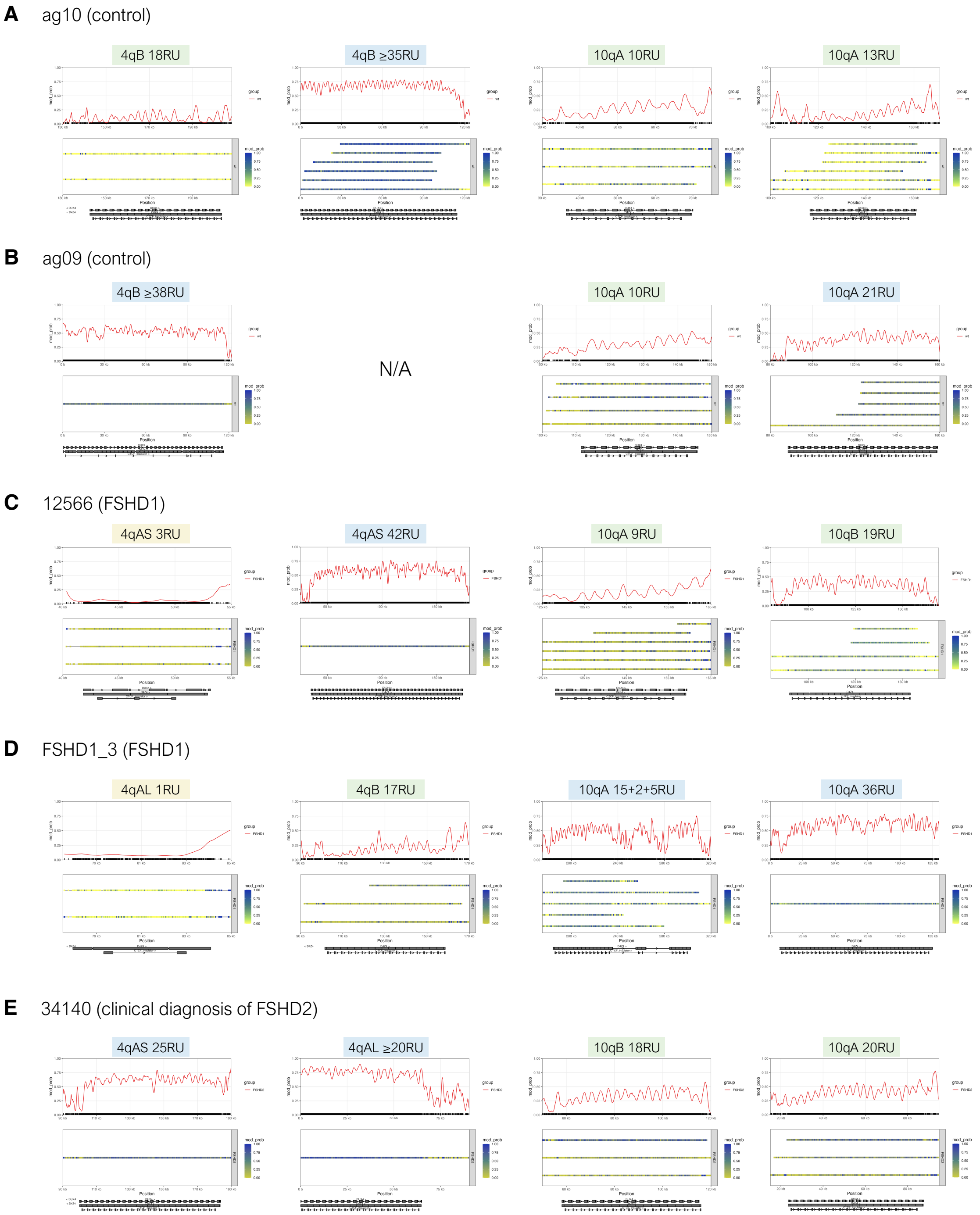


**Supplementary Figure S6. Allele-specific, array-wide D4Z4 methylation profiles for *SMCHD1*-wildtype fibroblast samples from whole-genome sequencing data**. Single-molecule and smoothed methylation plots for 4q and 10q D4Z4 alleles from (A) ag10: control sample (B) ag09: control sample (C) 12566: FSHD1 sample (D) FSHD1_3: FSHD1 sample (E) 34140: clinically diagnosed as FSHD2, yet lacking pathogenic variants in *SMCHD1*, *LRIF1*, and *DNMT3B*, as determined by Nanopore sequencing. Reads were obtained from ultra-long whole-genome Nanopore sequencing. Methylation plots were generated using NanoMethViz[20]. Annotations for D4Z4 repeat units, CTCF insulator regions, and *DUX4* exons are shown below each plot. Pathogenic 4qA alleles, ‘gray zone’ and intermediate-length alleles (8-20 repeat units), and long alleles (>20 repeat units) are shaded in yellow, green and blue, respectively.

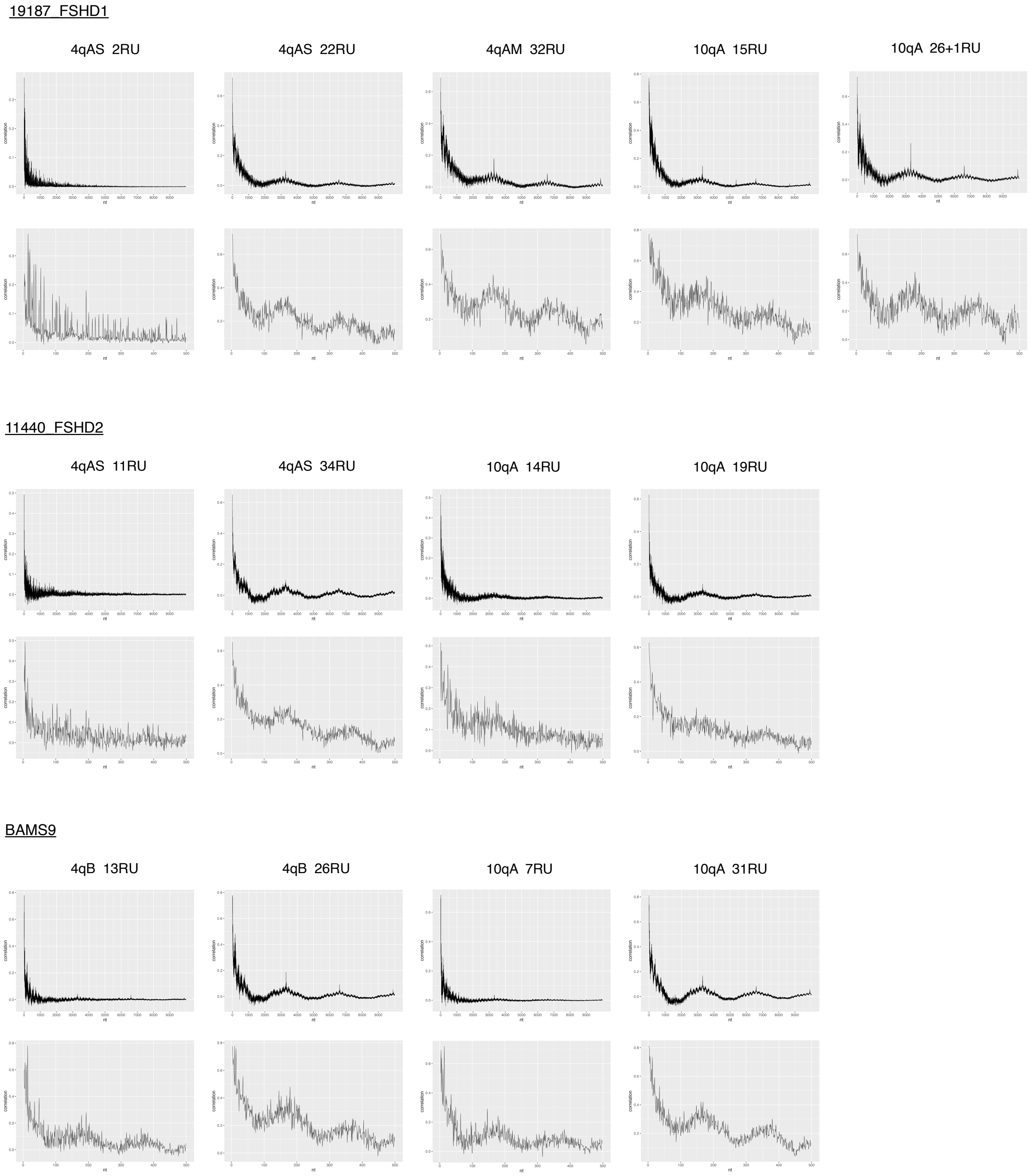


**Supplementary Figure S7. Autocorrelation of %5mC for CpG sites across the D4Z4 array for FSHD1, FSHD2 and BAMS alleles.** %5mC values are based on the output from modkit v0.2.5. Plots were generated for CpG sites separated by up to 10000nt (top panels) and up to 500nt (bottom panels), and show strong correlation for sites separated by ~3300nt and ~180nt, respectively.

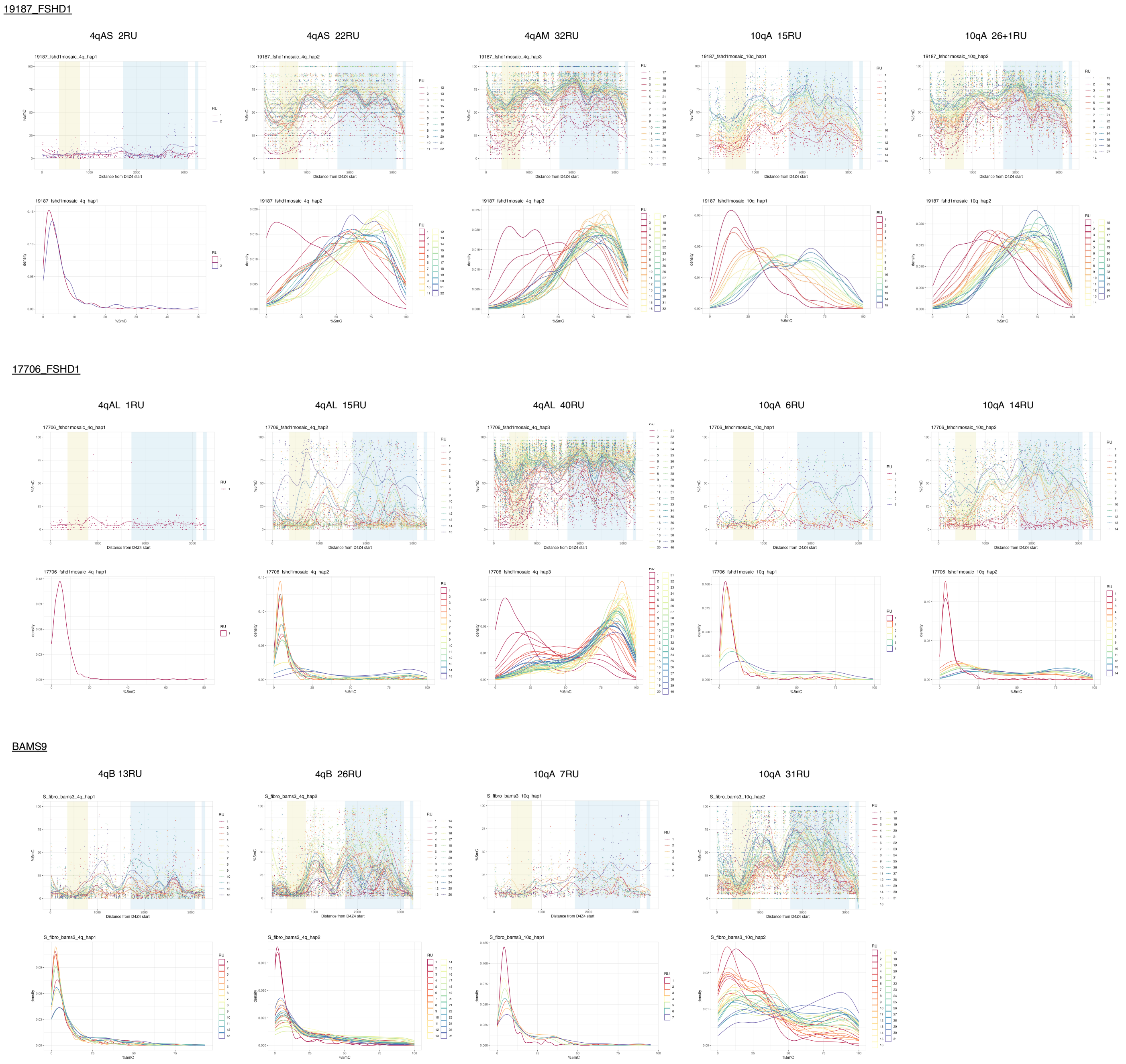


**Supplementary Figure S8. %5mC for CpG sites across individual D4Z4 units from FSHD1 and BAMS alleles.** The top row for each sample shows plots of the %5mC for each CpG site (individual dots) within each D4Z4 unit from the allele, alongside smoothed lines for each repeat unit. The locations of the CTCF insulator region and *DUX4* exons are shown in yellow and blue, respectively. The bottom row for each sample shows plots of the distributions of %5mC values for each D4Z4 from the allele. CpG sites with coverage <5 were filtered out before plotting.

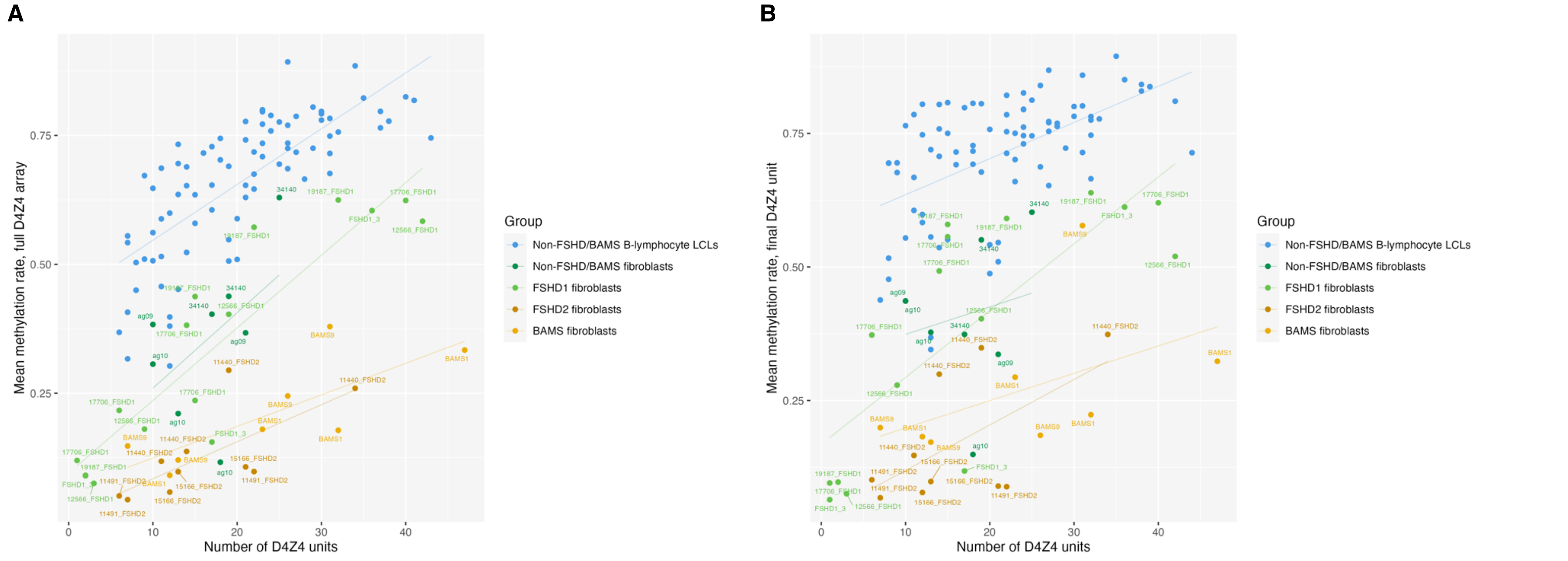


**Supplementary Figure S9**. **Correlation of overall D4Z4 methylation levels with the number of repeat units, grouped by cell type.** Mean methylation rates across (A) the full 4q/10q D4Z4 array or (B) the final D4Z4 unit against the number of D4Z4 units, plotted for alleles from FSHD1 fibroblasts, non-FSHD/BAMS (control) fibroblasts, FSHD2 fibroblasts, BAMS fibroblasts, and 30 non-FSHD/BAMS B-lymphocyte lymphoblastoid cell lines (LCLs) from the 1000 Genomes Project[9,16]. Mean methylation rate was calculated as the total number of methylated CpGs from all reads / the total number of methylated + unmethylated CpGs from all reads, across either the full D4Z4 array or the final D4Z4 unit. Regression lines were plotted for each group of samples. Sample 34140, while clinically-diagnosed as FSHD2, was grouped with 'non-FSHD/BAMS fibroblasts' due to the lack of pathogenic *SMCHD1, DNMT3B* or *LRIF1* variants and lack of FSHD- or BAMS-like hypomethylation.

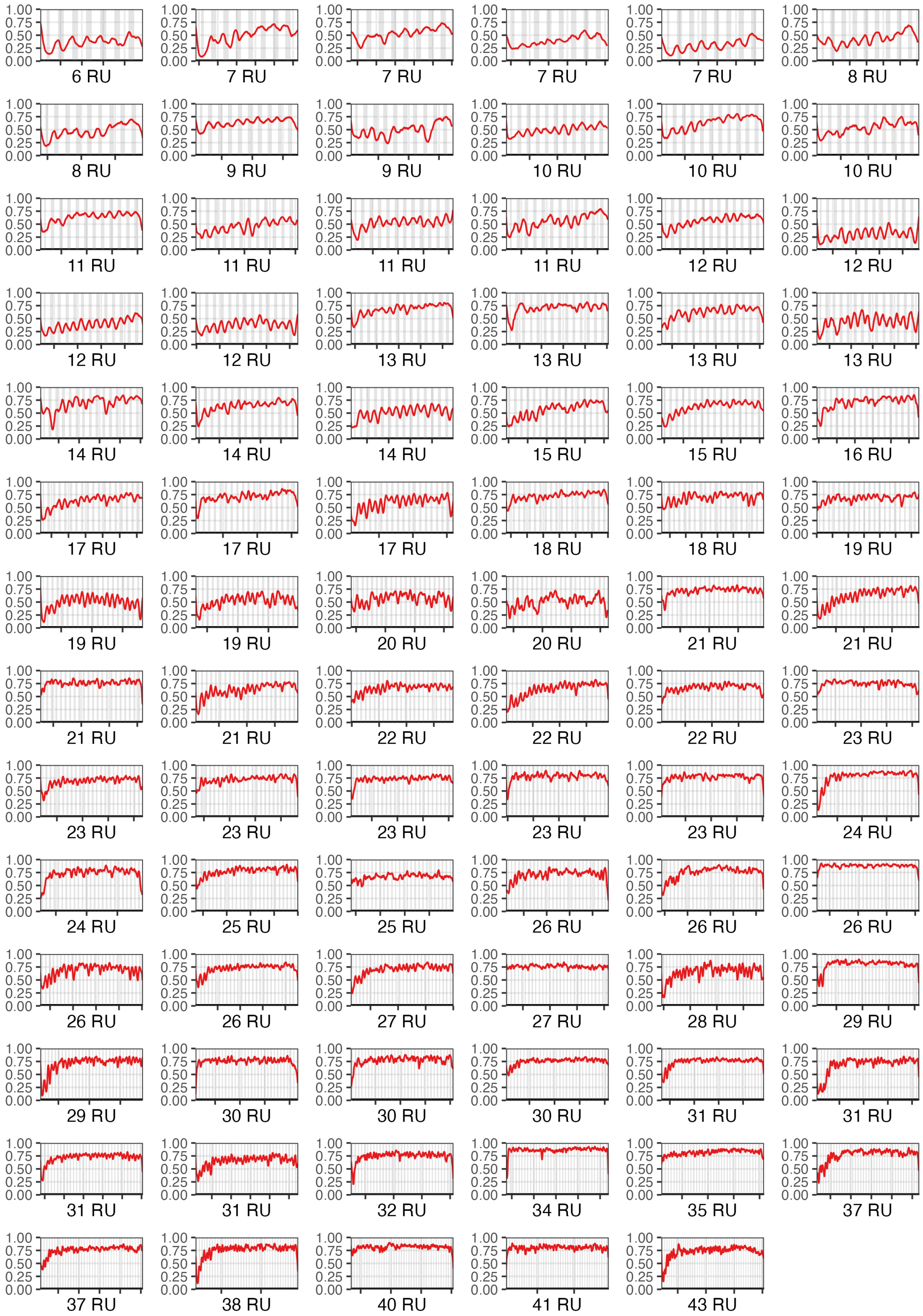


**Supplementary Figure S10. Smoothed methylation profiles for 4q and 10q D4Z4 alleles from B-lymphocyte lymphoblastoid cells lines (LCLs) from the 1000 Genomes Project[9,16].** Smoothed methylation plots were generated using NanoMethViz[20] in the range of 1.5kb upstream to 1.5kb downstream of the D4Z4 array; the y-axis represents the smoothed methylation probability based on the modification probability stored within the ML tag of the modBAM file. The location of *DUX4* exon 1 within each repeat unit is shown by gray vertical bars.

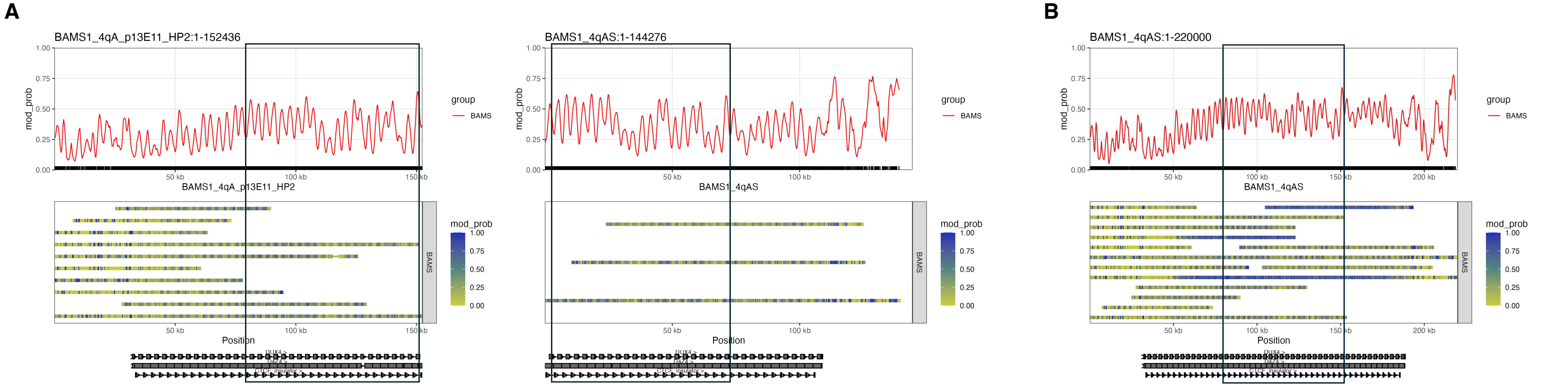


**Supplementary Figure S11. Genotype and methylation of the BAMS1 4qAS allele.** BAMS1 4qA reads overlapping the proximal end of the array were phased into two groups using WhatsHap[18], while 4qA reads overlapping the distal end of the array were phased based on AM/AS haplotype. (A) Smoothed methylation profile and single-molecule methylation plots for proximal D4Z4 reads (left) and distal D4Z4 reads (right) belonging to the BAMS1 4qAS allele, excluding hypermethylated reads. The presumed area of overlap between the proximal and distal reads (black rectangle) was determined based on the smoothed methylation profile. (B) Smoothed methylation profile and single-molecule methylation plots for proximal and distal D4Z4 4qAS reads aligned to a reconstructed full-length D4Z4 reference sequence with 47 repeat units. Four reads that mapped to the 4qAM haplotype had very high methylation levels compared to the other reads. The area of overlap between the proximal and distal reads is shown in the black rectangle.

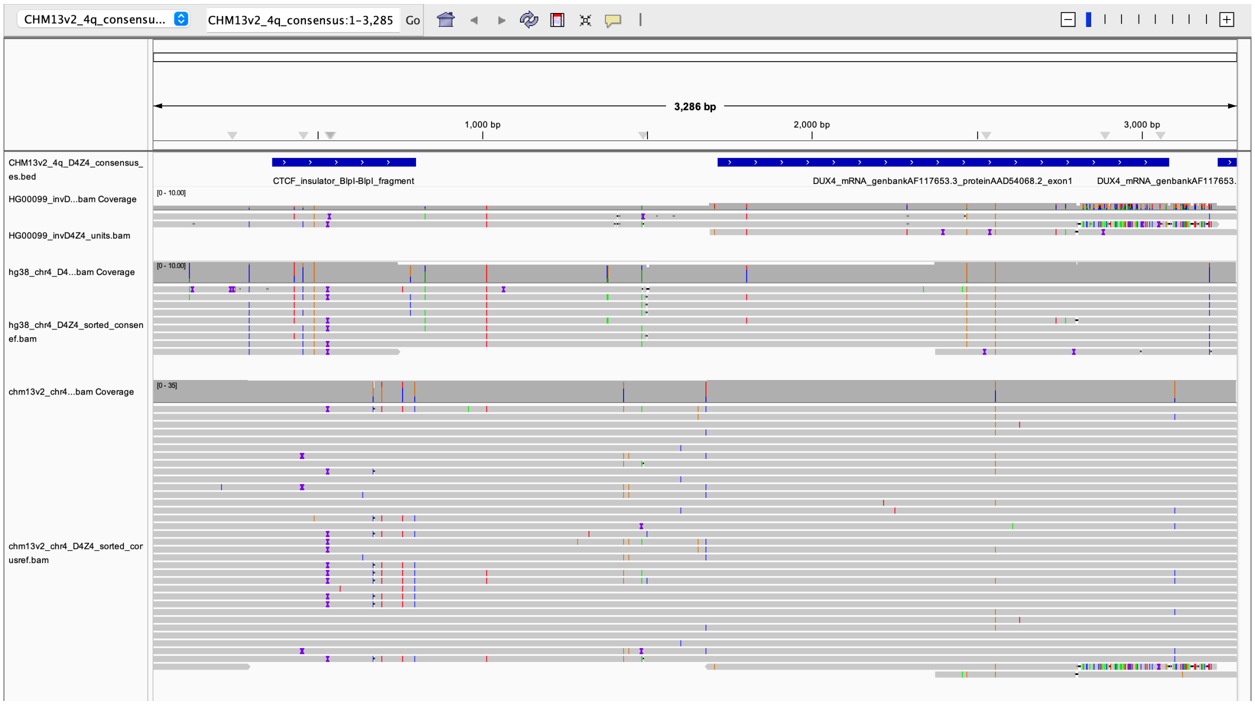


**Supplementary Figure S12. D4Z4 composition of the upstream inverted array on the HG00099 4qB allele.** Sequences of D4Z4 units were extracted from consensus sequences generated for the upstream inverted array using Racon[19], and aligned against a 4qA-type D4Z4 reference sequence (top track). Alignments of D4Z4 units from the GRCh38 4qB array (middle track) and the CHM13v2.0 4qA array (bottom track) are also shown, showing that the first (partial) and second (full) D4Z4 units of the upstream inverted array are 4qB-type, while the second half of the third (full) D4Z4 unit of the upstream inverted array corresponds to the normal sequence for the D4S2463 unit.
